# Systematic Review of Cerebral Phenotypes Associated with Monogenic Cerebral Small Vessel Disease

**DOI:** 10.1101/2021.11.12.21266276

**Authors:** Ed Whittaker, Sophie Thrippleton, Liza YW Chong, Victoria C Collins, Amy C Ferguson, David E Henshall, Emily Lancastle, Tim Wilkinson, Blair Wilson, Kirsty Wilson, Cathie Sudlow, Joanna Wardlaw, Kristiina Rannikmäe

## Abstract

**Background:** Cerebral small vessel disease (cSVD) is an important cause of stroke and vascular dementia. Most cases are multifactorial, but an emerging minority have a monogenic cause. While *NOTCH3* is the best-known gene, several others have been reported. We aimed to summarise the cerebral phenotypes associated with these more recent cSVD genes.

**Methods:** We performed a systematic review (PROSPERO: CRD42020196720), searching Medline/Embase (conception to July 2020) for any language publications describing *COL4A1/2, TREX1, HTRA1, ADA2*, or *CTSA* pathogenic variant carriers. We extracted data about individuals’ characteristics, clinical and vascular radiological cerebral phenotypes. We summarised phenotype frequencies per gene, comparing patterns across genes.

**Results:** We screened 6,485 publications including 402, and extracted data on 390 *COL4A1*, 123 *TREX1*, 44 *HTRA1* homozygous, 41 *COL4A2*, 346 *ADA2*, 82 *HTRA1* heterozygous, and 14 *CTSA* individuals. Mean age ranged from 15 (*ADA2*) to 59 years (*HTRA1* heterozygotes). Clinical phenotype frequencies varied widely: stroke 9% (*TREX1*) to 52% (*HTRA1* heterozygotes), cognitive features 0% (*ADA2*) to 64% (*HTRA1* homozygotes), psychiatric features 0% (*COL4A2*; *ADA2*) to 57% (*CTSA*). Among individuals with neuroimaging, vascular radiological phenotypes appeared common, ranging from 62% (*ADA2*) to 100% (*HTRA1* homozygotes; *CTSA*). White matter lesions were the most common pathology, except in *ADA2* and *COL4A2* cases, where ischaemic and haemorrhagic lesions dominated, respectively.

**Conclusions:** There appear to be differences in cerebral manifestations across cSVD genes. Vascular radiological changes were more common than clinical neurological phenotypes, and present in the majority of individuals with reported neuroimaging. However, these results may be affected by age and biases inherent to case reports. In the future, better characterisation of associated phenotypes, as well as insights from population-based studies, should improve our understanding of monogenic cSVD to inform genetic testing, guide clinical management, and help unravel underlying disease mechanisms.

## Introduction

Cerebral small vessel disease (cSVD) is recognised as an important cause of stroke and vascular cognitive impairment worldwide. The term cSVD describes a group of pathological processes which affect the small arteries, arterioles, venules, and capillaries within the brain (1). Features of cSVD on neuroimaging include subcortical infarcts, white matter lesions (WML), deep intracerebral haemorrhage (ICH), enlarged perivascular spaces (PVS), cerebral microbleeds and brain atrophy (2). Despite the increase in cSVD burden amongst an ageing population, the underlying disease mechanisms are incompletely understood, and therapeutic options limited, with vascular risk factor management remaining the mainstay of cSVD prevention and treatment (3).

While the majority of cSVD cases are thought to result from the interaction of multiple genetic variants and environmental factors, an important minority of cases are monogenic, i.e., caused by a pathogenic rare variant in a single gene. *NOTCH3* is the best known of these genes and is implicated in cerebral autosomal dominant arteriopathy with subcortical infarcts and leukoencephalopathy (CADASIL) (4). However, since *NOTCH3* was first described in 1996, several additional cSVD genes have been identified, including *COL4A1, TREX1, HTRA1, COL4A2, ADA2* and, most recently, *CTSA*. Pathogenic rare variants in these genes have been associated with various clinical phenotypes alongside cSVD, including extra-cerebral manifestations, as well as certain radiological features seen on neuroimaging (5).

Better characterisation of these rare disorders, including which radiological and clinical phenotypes are associated with specific genes, can inform genetic testing and counselling, including the appropriate selection of patients and screening of family members. This knowledge can also aid in the management of affected individuals, for example by guiding appropriate screening for certain associated phenotypes. Furthermore, an improved understanding of monogenic cSVD may offer insights into the disease mechanisms underlying sporadic cSVD, as there is increasing evidence to suggest an overlap of disease pathways involved in both sporadic and monogenic disease (6-8). Observations from large-scale genetic association studies have also shown common variation in monogenic cSVD genes to be associated with sporadic cSVD. Examples include *COL4A2* single nucleotide polymorphisms’ (SNPs) association with lacunar ischemic stroke and deep ICH, *HTRA1* SNPs association with ischaemic stroke, and possibly association of *NOTCH3* SNPs with WMLs (9-12).

We undertook a systematic literature review with the aim of identifying all reported individuals with putative pathogenic rare variants in any of the following monogenic cSVD genes: *COL4A1, TREX1, HTRA1, COL4A2, ADA2* and *CTSA*. We aimed to summarise and compare both clinical and vascular radiological cerebral phenotypes associated with each monogenic cSVD gene.

## Methods

### Registration

We have registered a PROSPERO protocol (ID: CRD42020196720) at https://www.crd.york.ac.uk/prospero/display_record.php?ID=CRD42020196720 (13). We followed the PRISMA guidelines (14).

### Search Strategy

We searched the MEDLINE and EMBASE databases using OvidSP (from conception to July 2020) for publications about individuals with pathogenic rare variants in any of our genes of interest: *COL4A1, TREX1, HTRA1, COL4A2, ADA2* or *CTSA*. We did not restrict the search by language or publication date; we limited it to human studies; we included conference abstracts. We used a previously-published search strategy (see Data Supplement) (5). In summary, the search included:

⍰ Text words, phrases, and Medical Subject Headings (MeSH) for relevant monogenic syndromes/diseases associated with our genes of interest, and
⍰ Text words, phrases, and MeSH terms associated with cSVD combined with those for our genes of interest and their proteins.

### Screening

We carried out the screening using Covidence (www.covidence.org). At least two reviewers (EW, ST, LYWC, DEH, BW, KR) independently screened titles and abstracts of all publications identified in our search, blinded to each other’s decisions. Full texts of studies included at this stage were then retrieved and screened by two reviewers for eligibility, recording any reasons for exclusion. We resolved disagreements through discussion and mutual consensus with a third reviewer. The included publications were combined with those identified via a previous systematic review (5).

### Inclusion / Exclusion Criteria

We included studies which met the following conditions:

- A case report, case series or other study design (except review papers) describing the clinical or cerebral radiological phenotype of ≥1 individual. Such description could be anything between stating that the individual was healthy, through to an in-depth case report.
- Genetically confirmed rare variant (in a heterozygous [HetZ], homozygous or compound heterozygous state [HomZ]) in any of our genes-of-interest.
- Study authors considered the rare variant to be probably or definitely pathogenic

We excluded studies describing individuals with rare variants in *CTSA* and *TREX1* associated with Galactosialidosis, Aicardi-Goutieres syndrome and chilblain or systemic lupus. We excluded individuals with a presumed pathogenic variant in >1 gene.

### Data extraction

From each included publication, we (one of EW, ST, LYWC, VC, EL, DEH, KR) extracted data on the first author, publication year, journal, and number of eligible individuals and pedigrees. For foreign language articles, we sought a full translation where an English language abstract did not provide sufficient information or was not available. For each eligible individual, we extracted data using a standardised form, including:

⍰ individual’s characteristics (region of origin, sex, age at time of assessment); genetic variant and resulting protein change;
⍰ clinical cerebral phenotype (presence, type and age at diagnosis of clinical stroke(s), cognitive features, psychiatric features, and headache);
⍰ vascular radiological cerebral phenotype (presence, location, burden, scan type used and age at diagnosis of ischaemia, ICH, WMLs, microbleeds, atrophy, enlarged PVS, calcification, and cerebral aneuryms; and
⍰ vascular risk factors (presence of one or more of hypertension, smoking, diabetes mellitus, excess alcohol consumption, or hypercholesterolaemia).

We selected the list of clinical cerebral phenotypes to extract to represent known manifestations of cSVD, including stroke, and the broad categories of cognitive and psychiatric features. We additionally included headache as phenotype-of-interest because of its association with several monogenic cSVD genes in OMIM (*ADA2, COL4A1, TREX1, HTRA1*). Finally, we also noted any other cerebral clinical phenotypes on our data extraction form.

We selected the list of vascular radiological cerebral phenotypes to extract to represent known manifestations of cSVD and again noted any other features on our data extraction form. Finally, we noted any specific radiological patterns to lesion location or severity that might help identify cases in everyday clinical practice.

To assess agreement in data extraction, at least 2 members of the team extracted data from 10% of publications, working independently, and blinded to each other’s decisions.

Where radiological imaging findings were described, the terminology used across publications varied widely, as has been noted previously in the literature (2). We made an effort to sort the imaging descriptions into our pre-specified categories to deal with the variable terminology (see Supplemental Methods for a list of decisions and assumptions), discussing uncertainties with an expert neuroradiologist (JW).

### Data synthesis

For each gene, we summarised the total number of relevant publications, pedigrees, individuals and rare variants, and the individuals’ characteristics. We summarised data on the presence or absence of each cerebral phenotype (clinical and vascular radiological) as well as cumulative evidence of any vascular radiological feature, to assess their apparent frequency. We compared findings between genes, highlighting shared patterns and differences in the frequencies of associated phenotypes.

We stratified the presence of clinical stroke and any vascular feature(s) on neuroimaging by presence of one or more vascular risk factors. We used the Chi-squared test (significance threshold of 0.05) to assess differences in phenotype frequency in patients with and without vascular risk factors.

### Variant pathogenicity assessment

We used the Ensembl Variant Effect Predictor (VEP) (15) to assess the consequences of the genetic variants included in our systematic review. We extracted information on the variants based on the following VEP sub-components: (i) SnpEff variant annotation and effect prediction tool to assess variant impact (16); (ii) ClinVar to assess variant’s clinical significance (17); (iii) SIFT to predict whether an amino acid substitution is likely to affect protein function (18); and, (iv) PolyPhen-2 to predict the effect of an amino acid substitution on the structure and function of a protein (19). Where conflicting evidence was provided for the same variant (usually because an allele may have a different effect in different transcripts), we selected the category with a more significant / negative effect. We calculated the results (expressed as percentages) among variants per each individual VEP sub-component.

## Results

We included 402 publications from 6485 identified for screening (Figure 1, Supplementary References). As in our previous systematic review (5), despite only being first reported in 2013, *ADA2* had the largest number of eligible publications (n=149), while the number of publications for other genes appears to be related to their order of discovery (*COL4A1* n=137; *TREX1* n=38; *HTRA1*^HomZ^ n=32; *COL4A2* n=20; *HTRA1*^HetZ^ n=32; *CTSA* n=5) (Figure 2). A likely explanation is the combination of existing treatment options and the severe early-onset systemic phenotype of *ADA2*, prompting more widespread genetic testing. We extracted data on 1,040 individuals, with the number of individuals per gene ranging from 14 (*CTSA*) to 390 (*COL4A1*), and the number of pedigrees ranging from 3 (*CTSA*) to 266 (*ADA2*). The percentage of pedigrees carrying a private variant ranged from 0% (*CTSA*) to 76% (*COL4A2*). As expected, the proportion carrying a private variant has decreased since our previous systematic review (5), presumably due to new reported individuals now becoming increasingly likely to have had their rare variant already identified previously (Figure 2).

**Figure 1:**
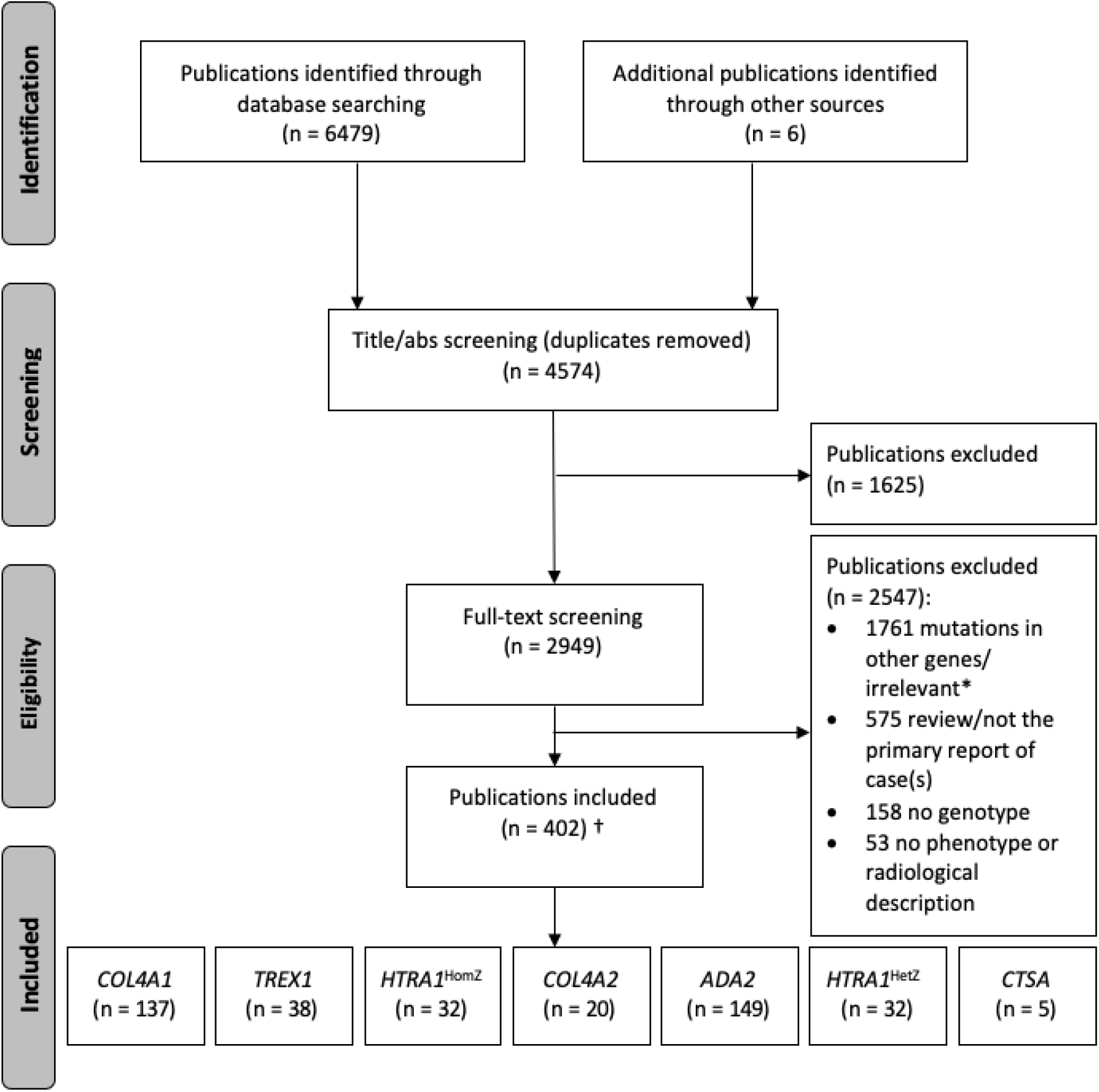
Selection of included publications. HomZ=homozygous/compound heterozygous; HetZ=heterozygous; n=number; abs=abstract. ^*^We identified *NOTCH3, FOXC1* and *PITX2* individuals as part of another systematic review. †1 publication reported both *HTRA1*^HomZ^ and *HTRA1*^HetZ^ individuals, 7 publications reported both *COL4A1/2* individuals, and 1 publication reported *HTRA1*^HetZ^, *COL4A1/2* and *TREX1* individuals, so the number of unique publications (402) is not the sum of publications per gene (413).

**Figure 2:**
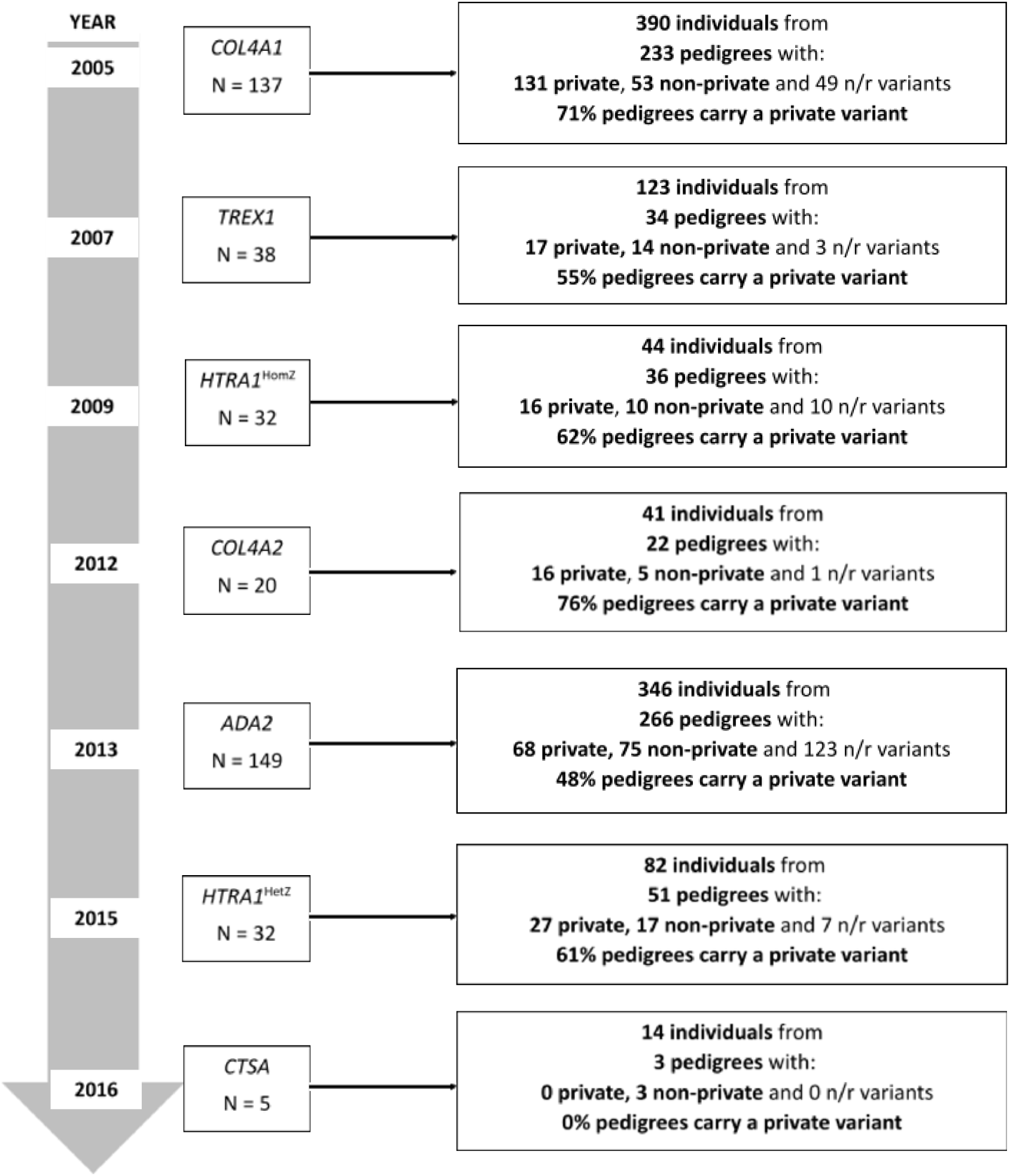
Number of included individuals and pedigrees. HomZ=homozygous/compound heterozygous; HetZ=heterozygous; N=number of publications; year=year gene first reported to be associated with cSVD; n/r=not reported. This figure is reporting on DNA change, variant was considered n/r where DNA change was not reported. For compound heterozygotes, if either variant was private, the pedigree was considered to carry a private variant. Where publications had not clearly reported these data (e.g. reporting 5 individuals with pathogenic *COL4A1* variants, but not specifying the variants, could refer to 5 individuals all carrying the same variant or each carrying a private variant), we assumed the maximum number of private variants (e.g. 5 private variants in this example).

The subset of included studies with data independently extracted for comparison showed 96.3% agreement.

### Summary of Individual’s Characteristics

The most common region of origin was Europe for *COL4A1, TREX1, COL4A2* and *CTSA* individuals (67% [263/390], 57% [70/123], 49% [20/41] and 100% [14/14], respectively); Asia for *HTRA1*^HomZ^ and *HTRA1*^HetZ^ individuals (75% [33/44] and 56% [46/82]), and Turkey for *ADA2* individuals (28% [98/346]). The region of origin was unknown in 0% to 16% of individuals per gene.

Sex distribution was generally approximated equal (45% to 52% female) where the number of individuals per gene was considered sufficient to allow meaningful comparison (>100 individuals per gene).

Data about the age of individuals at the time of assessment was not available for >20% of *COL4A1/2* individuals. Mean (median) age ranged from 15 (13) years for *ADA2* individuals to 59 (60) years *HTRA1*^HetZ^ individuals. For *COL4A1/2* and *ADA2*, the median age of individuals was below 18, while the age ranges were very broad (ranging from <1 to 77, 72 and 76, respectively)(Table 1).

**Table 1.**
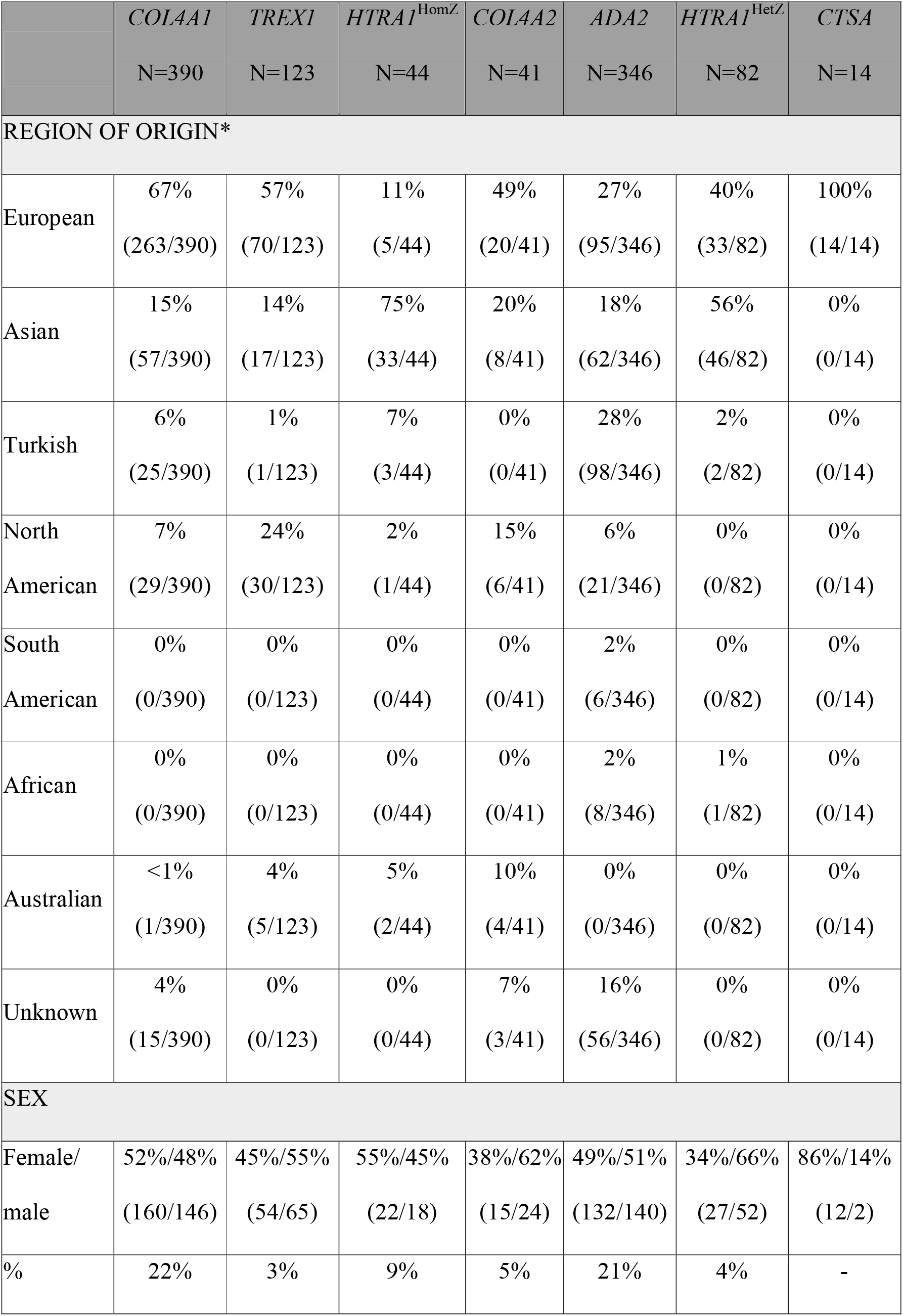
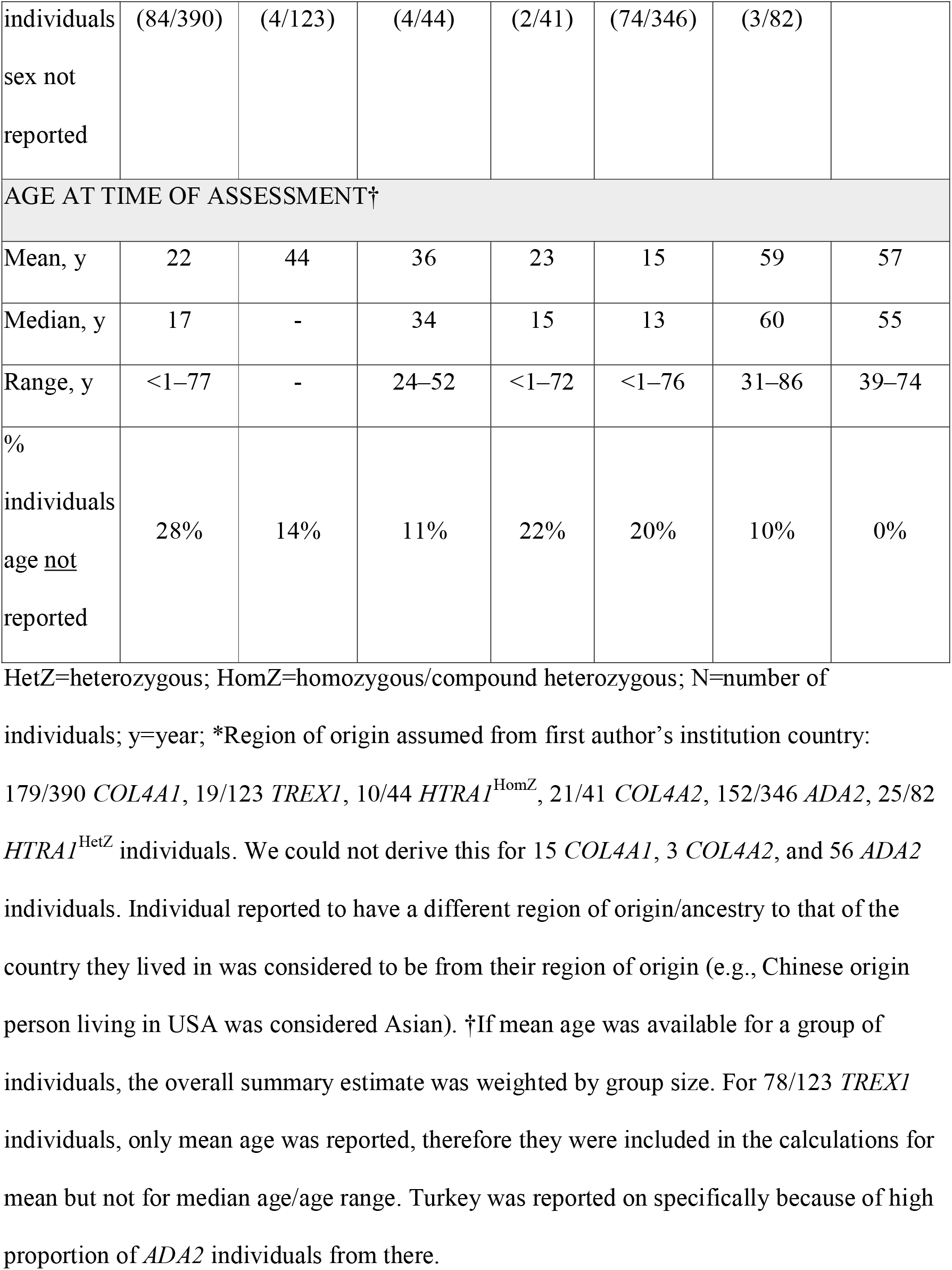
Summary of Case Characteristics.

### Frequency of clinical cerebral phenotypes

Cognitive features were the most common clinical cerebral phenotype for 4/7 genes (*HTRA1*^HomZ^, *COL4A2, HTRA1*^*H*etZ^, *CTSA*), stroke was the most common amongst *COL4A1* and *ADA2* individuals, and headache was most common amongst *TREX1* individuals (Figure 3, Supplementary Table 1).

**Figure 3.**
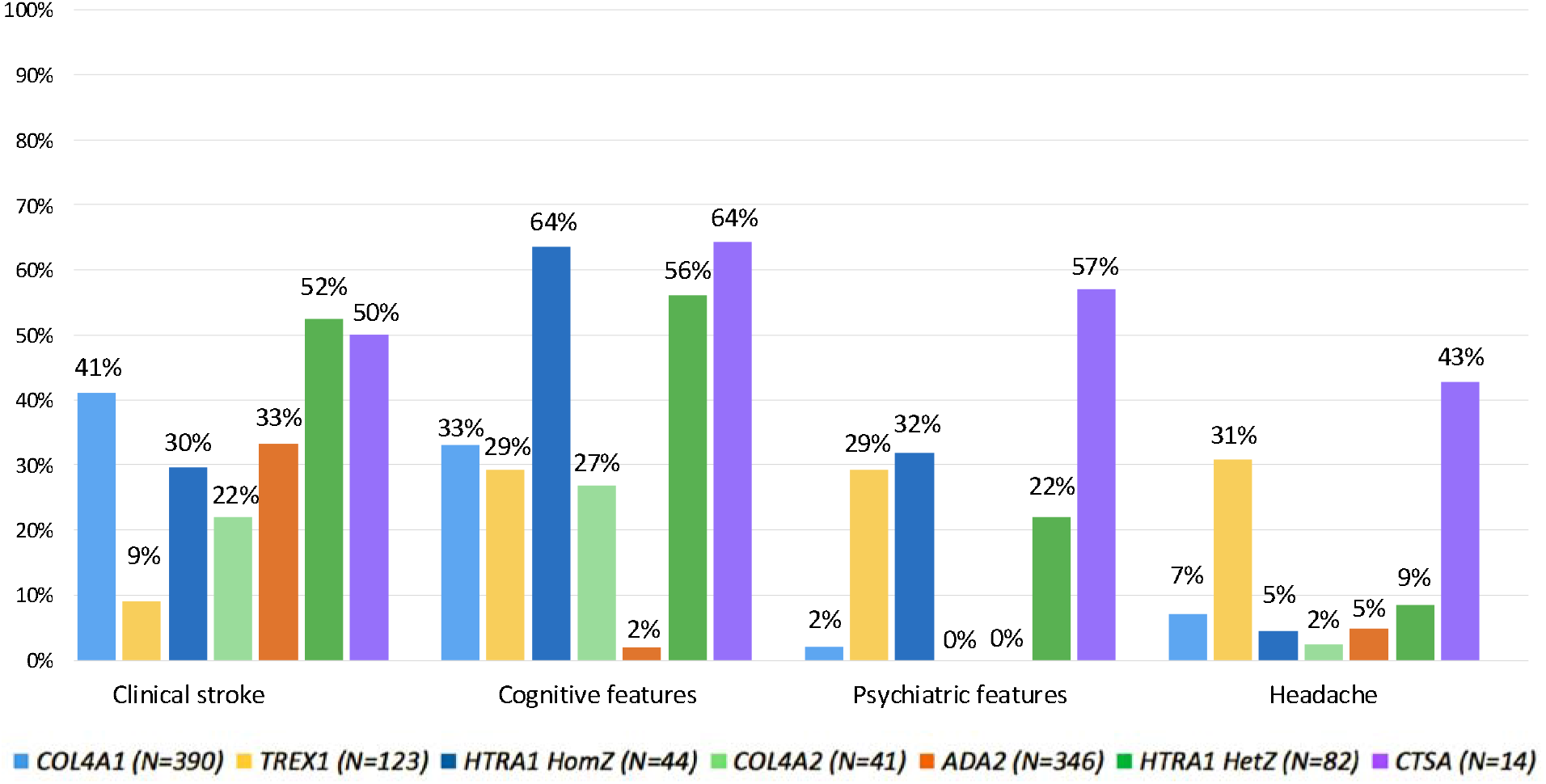
Frequency of Clinical Cerebral Phenotypes by Gene. HomZ=homozygous/compound heterozygous; HetZ=heterozygous;

#### Stroke

The frequency of clinical stroke ranged from 22% to 52% for 6/7 genes (*COL4A2* 22% [9/41]; *HTRA1*^HomZ^ 30% [13/44]; *ADA2* 33% [115/346]; *COL4A1* 41% [161/390]; *CTSA* 50% [7/14]; *HTRA1*^HetZ^ 52% [43/82]), while only 9% (11/123) of *TREX1* individuals were reported to have suffered a clinical stroke. Haemorrhagic events (ICH, porencephaly and intraventricular haemorrhage) were the most commonly reported stroke type amongst *COL4A1/2* individuals, affecting 73% (118/161) and 100% (9/9) of stroke cases, respectively. Ischaemic events (including arterial and venous ischaemic stroke, transient ischaemic attacks, and ocular vascular occlusions) were most common for all other genes and were reported in 54% to 100% of stroke cases (*HTRA1*^HomZ^ 54% [7/13]; *ADA2* 61% [70/115]; *HTRA1*^HetZ^ 62% [27/43]; *TREX1* 82% [9/11]; *CTSA* 100% [7/7]), although haemorrhagic events also occurred in a substantial minority.

#### Cognitive features

The frequency of cognitive features ranged from 27% to 64% for 6/7 genes (*COL4A2* 27% [11/41]; *TREX1* 29% [36/123]; *COL4A1* 33% [128/390]; *HTRA1*^HetZ^ 56% [46/82]; *HTRA1*^HomZ^ 64% [28/44]; and *CTSA* 64% [9/14]), while only 2% [7/346] of *ADA2* individuals were reported to have cognitive features. Developmental delay was present in over 80% of *COL4A1/2* individuals with cognitive features, however no cases of developmental delay were reported for other genes. For other genes, publications were generally lacking in detail so we could not draw conclusions about the nature and severity of cognitive decline (i.e., cognitive impairment vs dementia).

#### Psychiatric features

The frequency of psychiatric features ranged from 22% to 57% for 4/7 genes (*HTRA1*^*HetZ*^ 22% [18/82], *TREX1* 29% [36/124], *HTRA1*^*HomZ*^ *32% [14/44]*, and *CTSA* 57% [8/14], in ascending order of frequency). The most commonly reported psychiatric features were depression, followed by irritability and/or agitation. In contrast, only 2% (8/390) of *COL4A1* individuals reported psychiatric features, and no psychiatric features were reported amongst *COL4A2* and *ADA2* individuals (Supplementary Table I).

#### Headache

Headache was reported in 31% (38/123) of *TREX1* individuals and 43% (6/14) of *CTSA* individuals, with >80% of headache cases being specified as migraine. For all other genes, the frequency of headache ranged from 2% to 10%.

#### Other clinical cerebral phenotypes

32% *COL4A1/2* individuals (123/390 and 13/41, respectively) were reported to have suffered a seizure or have epilepsy. 43% (6/14) *CTSA* individuals were reported to suffer from vertigo and/or balance problems of unclear aetiology but suggested to signify brainstem and lower cranial nerve involvement.

### Frequency of radiological cerebral phenotypes

The proportion of individuals with neuroimaging (MRI, CT, MRA or CTA) was 74% (290/390) for *COL4A1*, 59% (73/123) for *TREX1*, 100% (44/44) for *HTRA1*^HomZ^, 76% (31/41) for *COL4A2*, 34% (119/346) for *ADA2*, 85% (70/82) for *HTRA1*^HetZ^ and 100% (14/14) for *CTSA*. Where neuroimaging was done, it included an MRI scan in 71% to 100% of cases. The rest of this section applies to those with neuroimaging only.

The majority of individuals showed vascular feature(s) on neuroimaging: ≥86% for all genes except *ADA2* (62%). Figure 4 shows the proportion of individuals with specific features suggestive of vascular brain disease, and Supplementary Table II shows the breakdown of these features by location and severity.

**Figure 4.**
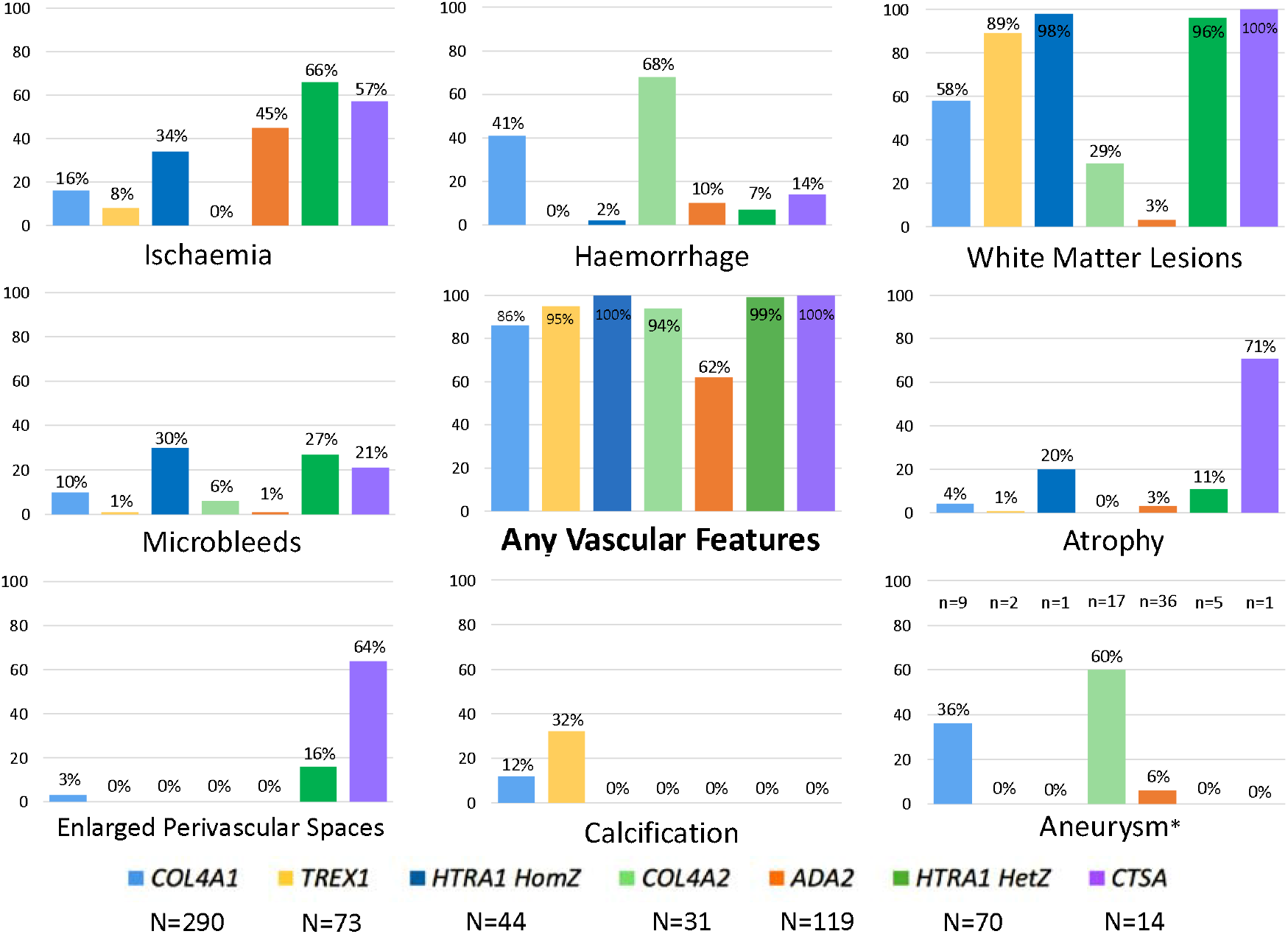
Frequency of Radiological Cerebral Phenotypes by Gene. HomZ=homozygous/compound heterozygous; HetZ=heterozygous; N=number of individuals with neuroimaging. Haemorrhage: intracerebral haemorrhage, intraventricular haemorrhage and/or porencephalic cysts.

#### Ischaemia

Presence ranged from 0% (*COL4A2*) to 66% (*HTRA1*^HetZ^). Ischaemia was the commonest radiological manifestation for *ADA2* individuals (45%). Location was reported for most individuals (80%), and as expected, where reported was mainly in deep/lacunar areas. Most individuals (70%) had multiple lesions.

#### ICH

Presence ranged from 0% (*TREX1*) to 68% (*COL4A2*). It was predominantly present in *COL4A1/2* individuals. However, ICH was also present in a small minority (7% to 10%) of *HTRA1, ADA2* and *CTSA* individuals. Porencephaly was present in *COL4A1/2* individuals only (61% and 76%, respectively) and intraventricular haemorrhage was present in *COL4A1* individuals only (7%). Location, where reported, was mostly deep. The burden is less clear: single lesions were common, though a minority of individuals did have multiple lesions.

#### WML

Presence ranged from 3% (*ADA2*) to 100% (*CTSA*). WMLs were the commonest radiological manifestation for 5/7 genes (not *COL4A2* and *ADA2*). Location was poorly reported, though where reported was common in the temporal regions in several genes. *CTSA* individuals appear to have lesions mainly in the frontal and parietal regions (though numbers are low). The burden of WML, where reported, was mostly severe, though the burden was not reported well (data missing for 51% individuals). The exception to this was *HTRA1*^HetZ^ individuals, who appear to have less severe WMLs. All *CTSA* individuals with WML with known location had temporal lobe sparing.

#### Microbleeds

Presence ranged from 1% (*TREX1*; *ADA2*) to 30% (*HTRA1*^HomZ^). Microbleeds were also common in *HTRA1*^HetZ^ individuals (27%). Location, where reported, was mostly deep. All individuals had multiple lesions where burden was reported.

#### Atrophy

Presence ranged from 0% (*COL4A2*) to 71% (*CTSA*). Location and burden were poorly described overall, and the low numbers make it difficult to make any conclusions.

#### Enlarged PVS

Presence was infrequent: enlarged PVS were present in *COL4A1* (3%), *HTRA1*^HetZ^ (16%) and *CTSA* (64%) individuals only.

#### Calcification

Presence was infrequent: calcification was present in *COL4A1/2* individuals only (12% and 32%, respectively).

#### Cerebral aneurysm

Present in 36% (13/36) *COL4A1*, 60% (3/5) *COL4A2* and 6% (1/17) *ADA2* individuals (of those with computed tomography angiograms or magnetic resonance angiograms reported).

#### Other radiological cerebral phenotypes

*COL4A1/2* individuals were also reported to manifest with schizencephaly (8% [24/290] *COL4A1* and 13% [4/31] *COL4A2* individuals) and cerebellar atrophy (5% [14/290] *COL4A1* and 3% [1/31] *COL4A2* individuals). 15% *TREX1* individuals (11/73) had pseudotumoral lesions.

#### Particular patterns to lesion location or severity to help identify cases in practice

A unique feature of *HTRA1*^*HomZ*^ individuals was the presence of arc-shaped hyperintense lesions from the pons to the middle cerebellar peduncles referred to as the ‘arc sign’ (9% [4/44] individuals) (20). A unique feature of *HTRA1*^*HetZ*^ individuals was the presence of dilated PVS in the basal ganglia referred to as ‘status cribrosum’ or ‘état crible’ (13% [9/70] individuals) (2, 21). Overall, the descriptions provided were not detailed enough to identify further patterns for other genes.

### Vascular Risk Factor Stratification

14% (134/928) of individuals across all genes were reported to have one or more vascular risk factors. Of these individuals, 62% (88/134) reported clinical stroke, compared to 34% (272/794) of individuals with no reported risk factors (p<0.01), while 78% (104/134) reported vascular features on neuroimaging, compared to 51% (401/794) of individuals with no reported risk factors (p<0.01) (Figure 5). The mean (median) age was 43 (48) years for those with one or more risk factor, and 22 (17) years for those with no reported risk factors. This analysis excludes individuals where data on risk factors and/or phenotypes was not available on an individual basis.

**Figure 5.**
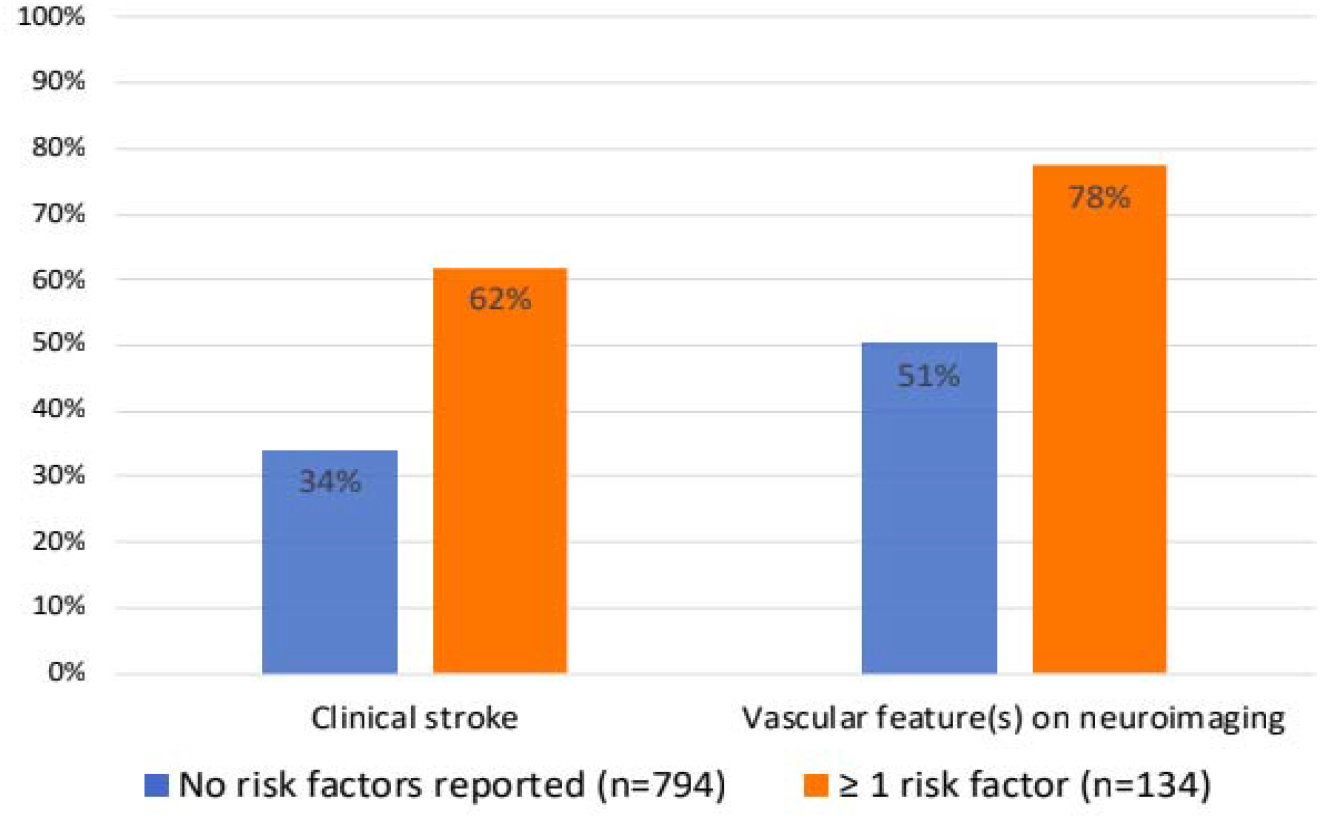
Frequency of Cerebral Phenotypes, stratified by presence of Vascular Risk Factors. N=number of individuals

### Variant pathogenicity assessment

VEP produced results from ≥1 of its sub-components for 15% to 66% of variants overall (SnpEff 66%, ClinVar 15%, SIFT 60%, and PolyPhen-2 62%), although there was substantial variability for these estimates across different genes. While the percentage of variants with supporting evidence of pathogenicity was high (81% to 99%) when studying only the group of variants with data available, this appeared much lower when including all variants regardless of whether or not VEP was able to process them (12% to 65%). Again, there was substantial variability across individual genes (Supplementary Table 3).

## Discussion

Vascular changes are commonly seen on neuroimaging in individuals with rare variant(s) in cSVD genes. Where data are available, the most frequent radiological manifestations are WMLs and ischaemic changes and, as expected, most lesions are deep. Common clinical phenotypes include clinical stroke, psychiatric symptoms and, most frequently reported, cognitive decline. Overall, radiological vascular phenotypes were more common than clinical neurological phenotypes. However, when interpreting these results, it is important to bear in mind that variation in the mean age of affected individuals may explain some of the differences in phenotypes between genes (e.g., increased age is a risk factor for both clinical stroke and vascular cerebral phenotypes on neuroimaging).

Both ICH and ischaemic stroke were described for all cSVD genes, although the most common stroke subtype was haemorrhagic for *COL4A1/2*, and ischaemic for the remaining genes. Enlarged perivascular spaces were infrequently reported, which may reflect this feature being less apparent with older imaging modalities, difficult to differentiate from other lesions such a lacunes (2), and/or less commonly reported on neuroimaging.

The frequency of both clinical stroke and vascular radiological features on neuroimaging was higher for those with at least one vascular risk factor, compared to those with no reported risk factors. However, vascular risk factors were generally poorly reported (therefore their presence cannot be excluded in most cases), age is highly likely to be a confounding factor, and individuals presenting with stroke/vascular radiological features are more likely to be investigated for vascular risk factors. More research is needed to understand the role for a focussed effort on addressing modifiable vascular risk factors in the management of monogenic cSVDs.

We identified only 14 individuals with a putative pathogenic variant in *CTSA*. This is likely (at least partly) explained by the relatively recent description of its association with cSVD, but the small overall number of affected individuals limit the conclusions that can be drawn about its phenotype associations.

The strengths of our study are: (1) a comprehensive search strategy, including foreign-language papers and abstracts; (2) systematic data extraction following a pre-set spreadsheet with a comprehensive list of variables to be collected, while also allowing for novel phenotypes to be recorded; (3) inclusion of several cSVD genes allowing comparisons to be made across these.

This research also has some limitations. Firstly, reporting for some variables was poor. For example, region of origin as a marker of ethnicity was frequently poorly reported and therefore often had to be assumed based on information such as the location of the authors’ institute. It is possible that some true differences between ethnicities may not have been revealed due to incorrect categorisation. The frequency of neuroimaging reporting was also low for some genes, and it is unknown if neuroimaging was not reported due to lack of positive findings, or whether it was not done at all. Secondly, case reports and case series have many inherent biases which are difficult to control for (e.g. testing bias, publication bias and reporting bias). In addition, the case reports included in this research appeared to lack use of a reporting structure. Current guidelines such as CARE (22, 23) don’t work so well in the field of rare genetic diseases and so new, tailored guidelines could help improve the consistency of reporting.

The frequency profile of clinical cerebral phenotypes associated with monogenic cSVDs suggests it is important to consider a broader spectrum of manifestations when identifying potential patients for genetic testing. Specifically, cognitive involvement appeared even more frequently than clinical stroke for several genes. Our results also show that in monogenic cSVD a radiological vascular phenotype is described more frequently than clinical cerebral phenotypes suggesting a potential benefit of radiological screening, both for patients and for at-risk family members.

It is also notable that across several monogenic cSVDs, WMLs were commonly identified in the temporal region, a feature which has previously been associated with CADASIL (caused by *NOTCH3* mutations) (24). It is therefore important to also consider other cSVD genes in the presence of this feature.

Finally, according to OMIM (https://www.omim.org), headache is a known phenotype associated with *TREX1* rare variants, thus its high frequency in *TREX1* individuals was expected. However other genes associated in OMIM with headache (*COL4A1, ADA2* and *HTRA1*) were not found to have a clear association with this phenotype in our review. 43% of *CTSA* individuals (albeit among a total of only 14 individuals) also reported headache, which is more than the expected population prevalence of 15% (25), suggesting a potentially novel associated phenotype. Epilepsy was another common phenotype in *COL4A1/2*, as suggested by OMIM and previous literature (26).

VEP predicted 81% to 99% of the processed variants to have a high likelihood of being pathogenic. However, since these percentages are calculated only among variants with data available, this introduces a bias, as some variants without data (e.g., synonymous SNPs) have a lower prior likelihood of being pathogenic. Adjusting these calculations to include all variants resulted in only 12% to 65% of variants having supporting evidence of pathogenicity, with substantial variability for results across individual genes. Also, it is possible that some variants have been submitted to ClinVar based on the same case-report/case-series included in our review. This makes it difficult to draw robust conclusions about included variants’ pathogenicity.

The findings summarised here have potential clinical implications for the diagnosis and follow up of monogenic cSVDs, especially in conjunction with previous data of associated extracerebral phenotypes (5). Having said this, to get a more comprehensive and less biased overview of the clinical and radiological consequences of monogenic cSVDs, further work should address these same questions using a genotype-first approach (i.e., studying this in a population-based setting and among individuals selected based on carrying the variant of interest, regardless of their phenotype). The emergence of prospective population-based studies with bio-samples yielding genetic data at scale, such as the UK Biobank (https://www.ukbiobank.ac.uk), will make this possible and complement our study findings.

In summary, we found that individuals with rare variant(s) in our genes of interest appear to develop vascular features on neuroimaging. Clinical stroke, cognitive and psychiatric features are also common. The phenotype profiles appear to differ across monogenic cSVD genes, however, these results may be affected by age and other biases inherent to case reports. In the future, better characterisation of associated phenotypes, as well as insights from population-based studies, should improve our understanding of monogenic cSVD to inform genetic testing, guide clinical management, and help unravel underlying disease mechanisms.

## Supporting information

Supplemental references

PRISMA checklist

## Data Availability

All data produced in the present work are contained in the supplemental material

## Acknowledgements

Mr Aidan Hutchison developed a database for data storage; Dr Michael Poon translated full-texts.

## Ethics

As a systematic review based on data from published studies this work does not require approval from an ethical standards committee.

