## Supplemental material for "Systematic Review of Cerebral Phenotypes Associated with Monogenic Cerebral Small Vessel Disease"

**Contents**

**Supplemental Methods**……………….…..………..………..………..………..………..………..…………..………….2

Decisions and Assumptions made when extracting data…………………..………..…………..……………2

**Supplemental Results……………………**……..………..………..………..………..………..…………...…..…...……3

Table I. Frequency and Subtypes of Cerebral Clinical Features..………..……………………..3

Table II. Frequency of Vascular Radiological Cerebral Phenotypes by Location and Severity …..….……..………..…………….………………………………………………………..……..………..…5

Table III. Variant Effect Predictor Output Summary……....………...………..…….…..……….10

Table IV. Variant Effect Predictor Outputs per Gene………………………………………....……12

A. *HTRA1……………*…..………..………..………..……….……..………..………..………....……12
B. *ADA2*….……..………..………..…...………..………..……….………..………..…………..……15
C. *COL4A1 ……………..*……..………..………..………..…....………..…..……..…………..……20
D. *COL4A2 ………………..*..………..………..………..………..………….………..……..…..……29
E. *TREX1……………..…………………………………………………………………………………….*31

**Appendix I.** Search Strategy…………..………..………..………..………..………....………..………….……...…32

**Supplemental Methods: Decisions and Assumptions made when extracting data**

**Demographic data**

- Age: sometimes specific ages weren’t reported but rather an approximate age or greater/less than a particular age was provided. In these cases we took a best estimation, erring towards overestimating age in some cases so as to minimise overestimation of the burden of the disease in younger brains. For example: <1 = 0, <2 = 1, <27 = 26, ≤26 = 26, early 50s = 52, mid-40s = 45.

**Clinical data**

- Clinical stroke classification required reporting of symptoms, i.e. not just radiological description
- Intellectual disability was classified under developmental delay

**Radiology data**

- When scan findings only described 'hemosiderin deposits' we did not take it to mean a confirmed bleed or microbleed
- Cerebral matter loss in <18 year old was recorded as ‘other’ rather than ‘atrophy’
- If a scan was described as showing 'stable findings'/’no changes’ or equivalent, we marked the scan as showing the same pathology as the previous scan of the same patient
- In general, author interpretations which used words such as ‘probable’ or ‘suggests’ were taken to mean the feature was present, while author interpretations which used words such as ‘possible’ or ‘might be’ were not sufficient to consider the feature present
- We took 'periventricular gliosis' to mean white matter lesions
- We classified haemorrhage at the splenium of corpus callosum as ‘deep’
- We took 'Hyperintense signal adjacent to the horn of the lateral ventricle' to mean periventricular white matter lesions
- External capsule, internal capsule, centrum semiovale and corona radiata locations qualified as deep
- Punctate hemorrhages were taken to mean brain microbleeds
- Regarding severity of white matter lesions, we assumed the following:
  - ‘Severe’ when described as: extensive, diffuse, severe, widespread, confluent, Fazekas score 3, disseminated
  - ‘Not severe’ when described as subtle, early/beginning confluent, limited, moderate, mild, weak, Fazekas score 1 or 2, punctiform
- If a scan was implied but not explicitly stated, we decided whether it was more likely a scan was done than not and assumed based on that – e.g. “haemorrhage in the right frontal area” was taken to mean a scan had been done
- We took a ‘petechial spot’ to mean a microbleed
- We took porencephalic cysts to be a subcategory of intracerebral haemorrhage

**Supplemental Results**

**Table I. Frequency and Subtypes of Cerebral Clinical Features**

|  |  | *COL4A1* (N=390) | *TREX1*  (N=123) | *HTRA1*^HomZ^ (N=44) | *COL4A2* (N=41) | *ADA2*  (N=346) | *HTRA1*^HetZ^  (N=82) | *CTSA* (N=14) |
| --- | --- | --- | --- | --- | --- | --- | --- | --- |
| **% (n/N)** | | | | | | | | |
| **Clinical stroke** | Unknown/ absent | 59 (229/390) | 91(112/123) | 70 (31/44) | 78 (32/41) | 67(231/346) | 48 (39/82) | 50 (7/14) |
|  | Present | 41 (161/390) | 9 (11/123) | 30 (13/44) | 22 (9/41) | 33 (115/346) | 52 (43/82) | 50 (7/14) |
|  | Ischaemic | 15 (24/161) | 82 (9/11) | 54 (7/13) | 0 (0/9) | 53 (61/115) | 53 (23/43) | 71 (5/7) |
|  | Ischaemic | 15 (24/161) | 73 (8/11) | 46 (6/13) | 11 (1/9) | 55 (63/115) | 44 (19/43) | 43 (3/7) |
|  | TIA | 2 (3/161) | 0 (0/11) | 8 (1/13) | 0 (0/9) | 5 (6/115) | 14 (6/43) | 43 (3/7) |
|  | Eye infarction | 0 (0/161) | 9 (1/11) | 0 (0/13) | 0 (0/9) | 3 (4/115) | 0 (0/43) | 14 (1/7) |
|  | Venous thrombosis/infarct | 0 (0/161) | 0 (0/11) | 0 (0/13) | 0 (0/9) | 0 (0/115) | 0 (0/43) | 14 (1/7) |
|  | Haemorrhagic | 72 (116/161) | 0 (0/11) | 8 (1/13) | 89 (8/9) | 12 (14/115) | 5 (2/43) | 0 (0/7) |
|  | ICH | 32 (51/161) | 0 (0/11) | 8 (1/13) | 22 (2/9) | 20 (23/115) | 14 (6/43) | 29 (2/7) |
|  | IVH | 4 (7/161) | 0 (0/11) | 0 (0/13) | 0 (0/9) | 0 (0/115) | 0 (0/43) | 0 (0/7) |
|  | Porencephalic cyst | 47 (76/161) | 0 (0/11) | 0 (0/13) | 78 (7/9) | 0 (0/115) | 0 (0/43) | 0 (0/7) |
|  | Ischaemic and haemorrhagic | 1 (2/161) | 0 (0/11) | 0 (0/13) | 11 (1/9) | 8 (9/115) | 9 (4/43) | 29 (2/7) |
|  | Unspecified/  no detail | 12 (19/161) | 18 (2/11) | 38 (5/13) | 0 (0/9) | 27 (31 /115) | 33 (14/43) | 0 (0/7) |
| **Cognitive features** | Unknown/ absent | 67 (262/390) | 71 (87/123) | 36 (16/44) | 73 (30/41) | 100(346/346) | 44 (36/82) | 36 (5/14) |
|  | Present | 33 (128/390) | 29 (36/123) | 64 (28/44)^#^ | 27 (11/41) | 0 (0/346) | 56 (46/82) | 64 (9/14) |
|  | Present (≥18 y) | 23 (30/131) | 34 (36/106) | 65 (20/31) | 0 (0/13) | 0 (0/85) | 62 (46/74) | 64 (9/14) |
|  | Dementia* | 3 (4/128)  17 (5/30) | 0 (0/36)  0 (0/36) | 32 (9/28)  45 (9/20) | 0 (0/11)  0 (0/0) | 0 (0/0)  0 (0/0) | 13 (6/46)  13 (6/46) | 0 (0/9)  0 (0/9) |
|  | Cognitive impairment- no ADL impact* | 2 (2/128)  7 (2/30) | 0 (0/36)  0 (0/36) | 0 (0/28)  0 (0/20) | 0 (0/11)  0 (0/0) | 0 (0/0)  0 (0/0) | 15 (7/46)  15 (7/46) | 0 (0/9)  0 (0/9) |
|  | Cognitive impairment- no ADL detail* | 12 (15/128)  (22/30) | 97 (35/36)  100 (35/36) | 68 (19/28)  55 (11/20) | 0 (0/11)  0 (0/0) | 0 (0/0)  0 (0/0) | 65 (30/46)  65 (30/46) | 100 (9/9)  100 (9/9) |
|  | Subjective cognitive decline* | 0 (0/128)  73 (0/30) | 0 (0/36)  0 (0/36) | 0 (0/28)  0 (0/20) | 0 (0/11)  0 (0/0) | 0 (0/0)  0 (0/0) | 7 (3/46)  7 (3/46) | 0 (0/9)  0 (0/9) |
|  | Developmental delay | 83 (106/128) | 0 (0/36) | 0 (0/28) | 100 (11/11) | 0 (0/0) | 0 (0/46) | 0 (0/9) |
| **Psychiatric features** | Unknown/ absent | 98 (382/390) | 71 (87/123) | 68 (30/44) | 100 (41/41) | 100(346/346) | 78 (64/82) | 43 (6/14) |
|  | Present | 2 (8/390) | 29 (36/123) | 32 (14/44) | 0 (0/41) | 0 (0/346) | 22 (18/82) | 57 (8/14) |
|  | Psychosis | 0 (0/8) | 6 (2/36) | 7 (1/14) | 0 (0/0) | 0 (0/0) | 6 (1/18) | 0 (0/8) |
|  | Depression symptoms | 25 (2/8) | 17 (6/36) | 64 (9/14) | 0 (0/0) | 0 (0/0) | 67 (12/18) | 88 (7/8) |
|  | Anxiety | 0 (0/8) | 3 (1/36) | 14 (2/14) | 0 (0/0) | 0 (0/0) | 0 (0/18) | 0 (0/8) |
|  | Irritability/ agitation | 25 (2/8) | 8 (3/36) | 64 (9/14) | 0 (0/0) | 0 (0/0) | 0 (0/18) | 13 (1/8) |
|  | Emotional lability | 13 (1/8) | 0 (0/36) | 21 (3/14) | 0 (0/0) | 0 (0/0) | 28 (5/18) | 13 (1/8) |
|  | OCD | 0 (0/8) | 0 (0/36) | 0 (0/14) | 0 (0/0) | 0 (0/0) | 6 (1/18) | 0 (0/8) |
|  | Unspecified/  no detail | 0 (0/8) | 78 (28/36) | 0 (0/14) | 0 (0/0) | 0 (0/0) | 0 (0/18) | 0 (0/8) |
| **Headache** | Unknown/ absent | 93 (362/390) | 69 (85/123) | 95 (42/44) | 98 (40/41) | 95 (329/346) | 91 (75/82) | 57 (8/14) |
|  | Present | 7 (28/390) | 31 (38/123) | 5 (2/44) | 2 (1/41) | 5 (17/346) | 9 (7/82) | 43 (6/14) |
|  | Migraine | 68 (19/28) | 84 (32/38) | 50 (1/2) | 100 (1/1) | 24 (4/17) | 43 (3/7) | 83 (5/6) |
|  | Unspecified | 32 (9/28) | 16 (6/38) | 50 (1/2) | 0 (0/1) | 76 (13/17) | 57 (4/7) | 17 (1/6) |

HetZ=heterozygous; HomZ=homozygous/compound heterozygous; N=overall number of individuals; n=number of affected individuals; ADL=activities of daily living; #8 cases with unknown age; * second row: only individuals ≥18 years; assumed Stam *et al* cohort were all ≥18 y.

**Table II. Frequency of Vascular Radiological Cerebral Phenotypes by Location and Severity**

|  | | | | ***COL4A1* (N=290)** | ***TREX1* (N=73)** | ***HTRA1*^HomZ^ (N=44)** | ***COL4A2* (N=31)** | ***ADA2* (N=119)** | ***HTRA1*^HetZ^**  **(N=70)** | ***CTSA* (N=14)** |
| --- | --- | --- | --- | --- | --- | --- | --- | --- | --- | --- |
| **% (n/N)** | | | | | | | | | | |
| **Ischaemia** | Totals | Present | | 16 (47/290) | 8 (6/73) | 34 (15/44) | 0 (0/31) | 44 (52/119) | 66 (46/70) | 57 (8/14) |
|  |  | Unknown/Absent | | 84(243/290) | 92 (67/73) | 66 (29/44) | 100(31/31) | 56 (67/119) | 34 (24/70) | 43 (6/14) |
|  | Location | Supratentorial | Deep/ lacunar | 43 (20/47) | 100 (6/6) | 53 (8/15) | 0 (0/0) | 42 (22/52) | 46 (21/46) | 75 (6/8) |
|  |  |  | Cortical | 2 (1/47) | 0 (0/6) | 0 (0/15) | 0 (0/0) | 2 (1/52) | 2 (1/46) | 25 (2/8) |
|  |  |  | Unknown | 4 (2/47) | 0 (0/6) | 20 (3/15) | 0 (0/0) | 10 (5/52) | 15 (7/46) | 0 (0/8) |
|  |  | Infratentorial | Brainstem | 51 (24/47) | 0 (0/6) | 53 (8/15) | 0 (0/0) | 44 (23/52) | 26 (12/46) | 0 (0/8) |
|  |  |  | Cerebellum | 2 (1/47) | 0 (0/6) | 0 (0/15) | 0 (0/0) | 2 (1/52) | 0 (0/46) | 25 (2/8) |
|  |  |  | Unknown | 0 (0/47) | 0 (0/6) | 0 (0/15) | 0 (0/0) | 0 (0/52) | 0 (0/46) | 0 (0/8) |
|  |  | Overall | Any deep | 83 (39/47) | 100 (6/6) | 67 (10/15) | 0 (0/0) | 77 (40/52) | 78 (36/46) | 100 (8/8) |
|  |  |  | No deep | 2 (1/47) | 0 (0/6) | 0 (0/15) | 0 (0/0) | 0 (0/52) | 0 (0/46) | 0 (0/8) |
|  |  |  | Unknown | 15 (7/47) | 0 (0/6) | 33 (5/15) | 0 (0/0) | 23 (12/52) | 22 (10/46) | 0 (0/8) |
|  | Burdenn | Single lesion | | 2 (1/47) | 33 (2/6) | 0 (0/15) | 0 (0/0) | 37 (19/52) | 0 (0/46) | 50 (4/8) |
|  |  | Multiple lesions | | 57 (27/47) | 50 (3/6) | 87 (13/15) | 0 (0/0) | 56 (29/52) | 100(46/46) | 38 (3/8) |
|  |  | Unknown | | 40 (19/47) | 17 (1/6) | 13 (2/15) | 0 (0/0) | 8 (4/52) | 0 (0/46) | 13 (1/8) |
| **Haemorrhage** | Totals | Present | | 41(118/290) | 0 (0/73) | 2 (1/44) | 68 (21/31) | 10 (12/119) | 7 (5/70) | 7 (1/14) |
|  |  | Unknown/Absent | | 59(172/290) | 100(73/73) | 98 (43/44) | 32 (10/31) | 90(107/119) | 93 (65/70) | 93(13/14) |
|  | Porencephaly | | | 61 (72/118) | 0 (0/0) | 0 (0/1) | 76 (16/21) | 0 (0/12) | 0 (0/5) | 0 (0/1) |
|  | IVH | | | 7 (8/118) | 0 (0/0) | 0 (0/1) | 0 (0/21) | 0 (0/12) | 0 (0/5) | 0 (0/1) |
|  | Location | Supratentorial | Deep/ lacunar | 25 (29/118) | 0 (0/0) | 0 (0/1) | 14 (3/21) | 50 (6/12) | 40 (2/5) | 100 (1/1) |
|  |  |  | Cortical | 2 (2/118) | 0 (0/0) | 0 (0/1) | 0 (0/21) | 8 (1/12) | 0 (0/5) | 0 (0/1) |
|  |  |  | Unknown | 13 (15/118) | 0 (0/0) | 0 (0/1) | 10 (2/21) | 42 (5/12) | 0 (0/5) | 0 (0/1) |
|  |  | Infratentorial | Brainstem | 2 (2/118) | 0 (0/0) | 0 (0/1) | 0 (0/21) | 0 (0/12) | 20 (1/5) | 0 (0/1) |
|  |  |  | Cerebellum | 6 (7/118) | 0 (0/0) | 100 (1/1) | 0 (0/21) | 0 (0/12) | 0 (0/5) | 0 (0/1) |
|  |  |  | Unknown | 0 (0/118) | 0 (0/0) | 0 (0/1) | 0 (0/21) | 0 (0/12) | 0 (0/5) | 0 (0/1) |
|  |  | Overall | Any deep | 56 (36/64) | 0 (0/0) | 100 (1/1) | 60 (3/5) | 50 (6/12) | 60 (3/5) | 100 (1/1) |
|  |  |  | No deep | 3 (2/64) | 0 (0/0) | 0 (0/1) | 0 (0/5) | 8 (1/12) | 0 (0/5) | 0 (0/1) |
|  |  |  | Unknown | 41 (26/64) | 0 (0/0) | 0 (0/1) | 40 (2/5) | 42 (5/12) | 40 (2/5) | 0 (0/1) |
|  | Burden | Single lesion | | 45 (53/118) | 0 (0/0) | 100 (1/1) | 76 (16/21) | 25 (3/12) | 100 (5/5) | 100 (1/1) |
|  |  | Multiple lesions | | 39 (46/118) | 0 (0/0) | 0 (0/1) | 19 (4/21) | 8 (1/12) | 0 (0/5) | 0 (0/1) |
|  |  | Unknown | | 16 (19/118) | 0 (0/0) | 0 (0/1) | 5 (1/21) | 67 (8/12) | 0 (0/5) | 0 (0/1) |
| **WML** | Totals | Present | | 58(167/290) | 89 (65/73) | 98 (43/44) | 29 (9/31) | 3 (3/119) | 96 (67/70) | 100(14/14) |
|  |  | Unknown/Absent | | 42(123/290) | 11 (8/73) | 2 (1/44) | 71 (22/31) | 97 116/119) | 4 (3/70) | 0(0/14) |
|  | Location | General | Periventricular only | 26 (43/167) | 9 (6/65) | 0 (0/43) | 78 (7/9) | 33 (1/3) | 7 (5/67) | 0 (0/14) |
|  |  |  | Deep only | 5 (9/167) | 2 (1/65) | 14 (6/43) | 0 (0/9) | 33 (1/3) | 24 (16/67) | 0 (0/14) |
|  |  |  | Both | 14 (24/167) | 2 (1/65) | 21 (9/43) | 0 (0/9) | 0 (0/3) | 25 (17/67) | 93 (13/14) |
|  |  |  | Unknown | 54 (91/167) | 88 (57/65) | 65 (28/43) | 22 (2/9) | 33 (1/3) | 43 (29/67) | 7 (1/14) |
|  |  | Region | Temporal | 7 (11/167) | 0 (0/65) | 30 (13/43) | 11 (1/9) | 0 (0/3) | 7 (5/67) | 0 (0/14) |
|  |  |  | Frontal | 3 (5/167) | 0 (0/65) | 5 (2/43) | 11 (1/9) | 0 (0/3) | 0 (0/67) | 86 (12/14) |
|  |  |  | Parietal | 2 (3/167) | 0 (0/65) | 2 (1/43) | 0 (0/9) | 0 (0/3) | 0 (0/67) | 86 (12/14) |
|  |  |  | Brainstem | 2 (3/167) | 0 (0/65) | 21 (9/43) | 0 (0/9) | 0 (0/3) | 9 (6/67) | 7 (1/14) |
|  |  |  | Unknown | 89(149/167) | 100(65/65) | 63 (27/43) | 89 (8/9) | 100 (3/3) | 85 (57/67) | 7 (1/14) |
|  | Burden | Severe | | 35 (59/167) | 5 (3/65) | 95 (41/43) | 22 (2/9) | 0 (0/3) | 12 (8/67) | 93 (13/14) |
|  |  | Not severe | | 12 (20/167) | 3 (2/65) | 0 (0/43) | 0 (0/9) | 0 (0/3) | 49 (33/67) | 0 (0/14) |
|  |  | Unknown | | 53 (88/167) | 92 (60/65) | 5 (2/43) | 78 (7/9) | 100 (3/3) | 39 (26/67) | 7 (1/14) |
| **Microbleeds** | Totals | Present | | 10 (29/290) | 1 (1/73) | 30 (13/44) | 6 (2/31) | 0 (0/119) | 27 (19/70) | 21 (3/14) |
|  |  | Unknown/Absent | | 90(261/290) | 99 (72/73) | 70 (31/44) | 94 (29/31) | 100(119/119) | 73 (51/70) | 79 (11/14) |
|  | Location | Supratentorial | Deep/ lacunar | 52 (15/29) | 0 (0/1) | 31 (4/13) | 50 (1/2) | 0 (0/0) | 47 (9/19) | 100 (3/3) |
|  |  |  | Cortical | 3 (1/29) | 0 (0/1) | 8 (1/13) | 0 (0/2) | 0 (0/0) | 0 (0/19) | 0 (0/3) |
|  |  |  | Unknown | 14 (4/29) | 0 (0/1) | 46 (6/13) | 0 (0/2) | 0 (0/0) | 26 (5/19) | 0 (0/3) |
|  |  | Infratentorial | Brainstem | 21 (6/29) | 0 (0/1) | 31 (4/13) | 0 (0/2) | 0 (0/0) | 16 (3/19) | 33 (1/3) |
|  |  |  | Cerebellum | 10 (3/29) | 0 (0/1) | 0 (0/13) | 0 (0/2) | 0 (0/0) | 11 (2/19) | 33 (1/3) |
|  |  |  | Unknown | 3 (1/29) | 0 (0/1) | 23 (3/13) | 0 (0/2) | 0 (0/0) | 0 (0/19) | 0 (0/3) |
|  |  | Overall | Any deep | 69 (20/29) | 0 (0/1) | 62 (8/13) | 50 (1/2) | 0 (0/0) | 53 (10/19) | 100 (3/3) |
|  |  |  | No deep | 0 (0/29) | 0 (0/1) | 0 (0/13) | 0 (0/2) | 0 (0/0) | 0 (0/19) | 0 (0/3) |
|  |  |  | Unknown | 31 (9/29) | 100 (1/1) | 38 (5/13) | 50 (1/2) | 0 (0/0) | 47 (9/19) | 0 (0/3) |
|  | Burden | Single lesion | | 14 (4/29) | 0 (0/1) | 0 (0/13) | 0 (0/2) | 0 (0/0) | 0 (0/19) | 33 (1/3) |
|  |  | Multiple lesions | | 76 (22/29) | 100 (1/1) | 85 (11/13) | 100 (2/2) | 0 (0/0) | 100 19/19) | 67 (2/3) |
|  |  | Unknown | | 10 (3/29) | 0 (0/1) | 15 (2/13) | 0 (0/2) | 0 (0/0) | 0 (0/19) | 0 (0/3) |
| **Cerebral atrophy** | Totals | Present | | 4 (12/290) | 1 (1/73) | 20 (9/44) | 0 (0/31) | 3 (4/119) | 11 (8/70) | 71 (10/14) |
|  |  | Unknown/Absent | | 96(278/290) | 99 (72/73) | 80 (35/44) | 100(31/31) | 97 (115/119) | 89 (62/70) | 29 (4/14) |
|  | Location | Global | | 25 (3/12) | 0 (0/1) | 0 (0/9) | 0 (0/0) | 25 (1/4) | 25 (2/8) | 0 (0/10) |
|  |  | Focal | | 42 (5/12) | 0 (0/1) | 11 (1/9) | 0 (0/0) | 25 (1/4) | 50(4/8) | 10 (1/10) |
|  |  | Unknown | | 33 (4/12) | 100 (1/1) | 89 (8/9) | 0 (0/0) | 50 (2/4) | 25 (2/8) | 90 (9/10) |
|  | Burden | Severe | | 42 (5/12) | 0 (0/1) | 0 (0/9) | 0 (0/0) | 0 (0/4) | 0 (0/8) | 0 (0/10) |
|  |  | Not severe | | 0 (0/12) | 100 (1/1) | 11 (1/9) | 0 (0/0) | 25 (1/4) | 50 (4/8) | 90 (9/10) |
|  |  | Unknown | | 58 (7/12) | 0 (0/1) | 89 (8/9) | 0 (0/0) | 75 (3/4) | 50 (4/8) | 10 (1/10) |
| **Calcification** | Totals | Present | | 12 (34/290) | 32 (23/73) | 0 (0/44) | 0 (0/31) | 0 (0/119) | 0 (0/70) | 0 (0/14) |
|  |  | Unknown/Absent | | 88(256/290) | 68 (50/73) | 100(44/44) | 100(31/31) | 100(119/119) | 100(70/70) | 100(14/14) |
| **Enlarged PVS** | Totals | Present | | 3 (8/290) | 0 (0/73) | 0 (0/44) | 0 (0/31) | 0 (0/119) | 16 (11/70) | 64 (9/14) |
|  |  | Unknown/Absent | | 97(282/290) | 100(73/73) | 100(44/44) | 100(31/31) | 100(119/119) | 84 (59/70) | 36 (5/14) |
| **Cerebral aneurysm** | Totals | Present | | 36 (13/36) | 0 (0/1) | 0 (0/9) | 60 (3/5) | 6 (1/17) | 0 (0/2) | 0 (0/1) |
|  |  | Unknown/Absent | | 64 (23/36) | 100 (1/1) | 100 (9/9) | 40 (2/5) | 94 (16/17) | 100 (2/2) | 100 (1/1) |

HetZ=heterozygous; HomZ=homozygous/compound heterozygous; N=overall number of individuals with neuroimaging; n=number of affected individuals; WML=white matter lesions(s); PVS=perivascular space(s);

**Table III. Variant Effect Predictor Output Summary**

| **Number of variants** | | | | | | | **% variants with info** | **% pathogenic* among variants with data** | **% pathogenic***  **among**  **all variants** |
| --- | --- | --- | --- | --- | --- | --- | --- | --- | --- |
| **Variant impact/classification of severity (SNPEff)** | | | | | | | | | |
|  | no info | low | moderate* | high* |  |  |  |  |  |
| *HTRA1* | 7 | 0 | 35 | 11 |  |  | 87% | 100% | 87% |
| *ADA2* | 43 | 3 | 24 | 18 |  |  | 51% | 93% | 48% |
| *COL4A1* | 43 | 0 | 88 | 23 |  |  | 72% | 100% | 72% |
| *COL4A2* | 1 | 0 | 14 | 1 |  |  | 94% | 100% | 94% |
| *TREX1* | 21 | 0 | 2 | 8 |  |  | 32% | 100% | 32% |
| *CTSA* | 1 | 0 | 0 | 0 |  |  | 0% | 0% | 0% |
| Total | 116 | 3 | 163 | 61 |  |  | 66% | 99% | 65% |
| **Clinical significance (ClinVar)** | | | | | | | | | |
|  | no info | uncertain clinical significance | benign | likely benign | likely pathogenic* | pathogenic* |  |  |  |
| *HTRA1* | 30 | 3 | 0 | 0 | 5 | 15 | 43% | 87% | 38% |
| *ADA2* | 76 | 2 | 1 | 0 | 6 | 3 | 14% | 75% | 10% |
| *COL4A1* | 150 | 1 | 0 | 0 | 0 | 3 | 3% | 75% | 2% |
| *COL4A2* | 5 | 0 | 0 | 3 | 3 | 5 | 69% | 73% | 50% |
| *TREX1* | 29 | 0 | 0 | 0 | 1 | 1 | 6% | 100% | 6% |
| *CTSA* | 1 | 0 | 0 | 0 | 0 | 0 | 0% | 0% | 0% |
| Total | 291 | 6 | 1 | 3 | 15 | 27 | 15% | 81% | 12% |
| **Impact on protein function (SIFT)** | | | | | | | | | |
|  | no info | tolerated | deleterious* |  |  |  |  |  |  |
| *HTRA1* | 18 | 1 | 34 |  |  |  | 66% | 97% | 64% |
| *ADA2* | 43 | 4 | 41 |  |  |  | 51% | 91% | 47% |
| *COL4A1* | 46 | 9 | 99 |  |  |  | 70% | 92% | 64% |
| *COL4A2* | 1 | 2 | 13 |  |  |  | 94% | 87% | 81% |
| *TREX1* | 29 | 1 | 1 |  |  |  | 6% | 50% | 3% |
| *CTSA* | 1 | 0 | 0 |  |  |  | 0% | 0% | 0% |
| Total | 138 | 17 | 188 |  |  |  | 60% | 92% | 55% |
| **Impact on protein structure and function (PolyPhen-2)** | | | | | | | | | |
|  | no info | benign | possibly damaging* | probably damaging* |  |  |  |  |  |
| *HTRA1* | 18 | 0 | 4 | 31 |  |  | 66% | 100% | 66% |
| *ADA2* | 43 | 5 | 1 | 39 |  |  | 51% | 89% | 45% |
| *COL4A1* | 40 | 2 | 15 | 97 |  |  | 74% | 98% | 73% |
| *COL4A2* | 1 | 0 | 4 | 11 |  |  | 94% | 100% | 94% |
| *TREX1* | 29 | 2 | 0 | 0 |  |  | 6% | 0% | 0% |
| *CTSA* | 1 | 0 | 0 | 0 |  |  | 0% | 0% | 0% |
| Total | 132 | 9 | 24 | 178 |  |  | 62% | 96% | 59% |

*category considered to provide supporting evidence for pathogenicity; SnpEff classifies each variant in one of the following output categories: high impact (variant is assumed to have a disruptive impact in the protein, probably causing protein truncation, loss of function or triggering nonsense mediated decay), moderate impact (non-disruptive variant that might change protein effectiveness), and low impact (variant assumed to be mostly harmless or unlikely to change protein behaviour). The ‘modifier’ category is taken to represent no information about these categories; ClinVar assigns each variant as pathogenic, likely pathogenic, likely benign, benign, or of uncertain clinical significance; SIFT predicts whether an amino acid substitution is likely to affect protein function based on sequence homology and the physico-chemical similarity between the alternate amino acids, concluding with a qualitative prediction if a variant is deleterious or tolerated; PolyPhen-2 predicts the effect of an amino acid substitution on the structure and function of a protein using sequence homology, 3D structures where available, and a number of other databases and tools. It classifies each variant as probably damaging, possibly damaging or benign.

**TABLE IV. Variant Effect Predictor outputs per gene**

**Table IV A. *HTRA1***

| **Genetic mutation** | **Protein change** | **Variant information** |
| --- | --- | --- |
| c.589C>T | p.R197X | Stop gained, likely deleterious, high impact. Pathogenic |
| c.865C>T | p.Q289X | Stop gained, likely deleterious, high impact. Pathogenic |
| c.1108C>T | p.R370X | Stop gained, high impact variant. Pathogenic/likely pathogenic |
| c.904C>T | p.R302X | Stop gained, high impact variant. Likely pathogenic |
| c.502A.T | p.K168ter | Stop gained, high impact variant |
| c.847G>T | p.G283Ter | Stop gained, high impact variant |
| c.983C>A | p.S328* | Stop gained, high impact variant |
| c.1005+1G>T |  | Splice donor variant, high impact |
| c.971A>C | p.N324T | Missense variant, possible splice region variant with moderate impact. Probably damaging to protein structure and conflicting evidence of tolerated/deleterious to protein function. Likely pathogenic |
| c.754G>A | p.A252T | Missense variant, non-coding exon variant with moderate impact. Possibly damaging to protein structure and deleterious to protein function. Pathogenic |
| c.956C>T | p.T319I | Missense variant, moderate impact. Probably damaging to protein structure and likely to have deleterious effect on protein function |
| c.451C>A | p.Q151K | Missense variant, moderate impact and potential modifier of both upstream and downstream gene regulation (ARMS2), and lncRNA. Probably damaging to protein structure and likely to have deleterious effect on protein function. Uncertain clinial significance |
| c.359G>A | p.G120D | Missense variant, moderate impact and potential modifier of both upstream and downstream gene regulation (ARMS2), and lncRNA. Probably damaging to protein structure and likely to have deleterious effect on protein function. Likely pathogenic |
| c.361A>C | p.S121R | Missense variant, moderate impact and potential modifier of both upstream and downstream gene regulation (ARMS2), and lncRNA. Probably damaging to protein structure and likely to have deleterious effect on protein function |
| c.397C>G | p.R133G | Missense variant, moderate impact and potential modifier of both upstream and downstream gene regulation (ARMS2), and lncRNA. Possibly damaging to protein structure but tolerated by protein function |
| c.367G>T | p.A123S | Missense variant, moderate impact and potential modifier of both upstream and downstream gene regulation (ARMS2), and lncRNA. Only possibly damaging to protein structure and likely to have deleterious effect on protein function |
| c.821G>A | p.R274Q | Missense variant with moderate impact. Probably/possibly damaging to protein structure and deleterious/some reports of tolerated to protein function. Pathogenic |
| c.496C>T | p.R166C | Missense variant with moderate impact. Probably/possibly damaging to protein structure and deleterious to protein function |
| c.517G>A | p.A173T | Missense variant with moderate impact. Probably/possibly damaging to protein structure and deleterious to protein function |
| c.517G>C | p.A173P | Missense variant with moderate impact. Probably/possibly damaging to protein structure and deleterious to protein function |
| c.856T>G | p.F286V | Missense variant with moderate impact. Probably/possibly damaging to protein structure and deleterious to protein function |
| c.854C>A | p.P285Q | Missense variant with moderate impact. Probably damaging to protein structure and deleterious to protein function. Uncertain clinical significance/pathogenic |
| c.854C>T | p.P285L | Missense variant with moderate impact. Probably damaging to protein structure and deleterious to protein function. Uncertain clinical significance/pathogenic |
| c.616G>A | p.G206R | Missense variant with moderate impact. Probably damaging to protein structure and deleterious to protein function. Uncertain clinical significance |
| c.961G>A | p.A321T | Missense variant with moderate impact. Probably damaging to protein structure and deleterious to protein function. Uncertain clinical significance |
| c.1091T>C | p.L364P | Missense variant with moderate impact. Probably damaging to protein structure and deleterious to protein function. Pathogenic |
| c.497G>T | p.R166L | Missense variant with moderate impact. Probably damaging to protein structure and deleterious to protein function. Pathogenic |
| c.614C>G | p.S205C | Missense variant with moderate impact. Probably damaging to protein structure and deleterious to protein function. Pathogenic |
| c.852C>A | p.S284R | Missense variant with moderate impact. Probably damaging to protein structure and deleterious to protein function. Pathogenic |
| c.883G>A | p.G295R | Missense variant with moderate impact. Probably damaging to protein structure and deleterious to protein function. Pathogenic |
| c.889G>A | p.V297M | Missense variant with moderate impact. Probably damaging to protein structure and deleterious to protein function. Pathogenic |
| c.536T>A | p.I179N | Missense variant with moderate impact. Probably damaging to protein structure and deleterious to protein function. Likely pathogenic |
| c.827G>C | p.G276A | Missense variant with moderate impact. Probably damaging to protein structure and deleterious to protein function. Likely pathogenic |
| c.1021G>A | p.G341J | Missense variant with moderate impact. Probably damaging to protein structure and deleterious to protein function. |
| c.524T>A | p. V175E | Missense variant with moderate impact. Probably damaging to protein structure and deleterious to protein function |
| c.527T>C | p.V176A | Missense variant with moderate impact. Probably damaging to protein structure and deleterious to protein function |
| c.646 G>A | p.V216 M | Missense variant with moderate impact. Probably damaging to protein structure and deleterious to protein function |
| c.847G>A | p.G283R | Missense variant with moderate impact. Probably damaging to protein structure and deleterious to protein function |
| c.848G>A | p.G283E | Missense variant with moderate impact. Probably damaging to protein structure and deleterious to protein function |
| c.850A>G | p.S284G | Missense variant with moderate impact. Probably damaging to protein structure and deleterious to protein function |
| c.905G>A | p.R302Q | Missense variant with moderate impact. Probably damaging to protein structure and deleterious to protein function |
| c.1348G>C | p.D450H | Missense variant with moderate impact. Only possibly damaging to protein structure and deleterious to protein function. |
| c.184-185del |  | Intronic variant, with possible impact on both upstream and downsteam gene regulation, ARMS2. Possible influence on lncRNA. |
| c.830_831delAG | p.E277Vfs | Intronic variant with possible influence on upstream gene |
| c.126delG | p.E42fs | Frameshift variant with high impact. Pathogenic |
| c.543delT | p.A182Pfs*33 | Frameshift mutation with high impact. Potentially leading to premature stop. Pathogenic |
| c.739delG | p.E247Rfs | Frameshift mutation with high impact. Potentially leading to premature stop |
| c.958G>A | p.D320N | Missense variant with moderate impact. Probably damaging to protein structure and deleterious to protein function. |

**Table IV B. *ADA2***

| **Genetic mutation** | **Protein change** | **Variant information** |
| --- | --- | --- |
| c.982G>A | p.E328K | Missense variant with potential impact on upstream and downstream gene regulation, possible 3'UTR variant involved in nonsense-mediated decay. Possible impact on processing of pseudogene, and CTCF binding site. Probably damaging/benign to protein structure and likely deleterious to protein function/potentially tolerated. |
| c.138/144delG |  | 5'UTR variant, intronic variant with possible impact on regulation of upstream gene through processing of pseudogene/nonsense mediated decay |
| c.37_39del | p.K13del | 5'UTR variant, intronic variant with possible impact on regulation of upstream gene through processing of pseudogene/nonsense mediated decay |
| c.143_144insG | p.R49Afs*13 | Frameshift variant with high impact, possible impact on both upstream and downstream gene regulation. Likely pathogenic |
| c.144 dup | p.R49Afs*13 | Frameshift variant with high impact, possible impact on both upstream and downstream gene regulation. Likely pathogenic |
| c.144_145ins |  | Frameshift variant with high impact, possible impact on both upstream and downstream gene regulation. Likely pathogenic |
| c.144del | p.R49Gfs*4 | Frameshift variant with high impact, possible impact on both upstream and downstream gene regulation. Likely pathogenic |
| c.144delG | p.R49fs | Frameshift variant with high impact, possible impact on both upstream and downstream gene regulation. Likely pathogenic |
| c.144dupG | p.R49fs | Frameshift variant with high impact, possible impact on both upstream and downstream gene regulation. Likely pathogenic |
| c.629delT |  | Frameshift variant, high impact with potential impact on both upstream and downstream genes. |
| c.427del | p.I143Sfs*41 | Frameshift variant, high impact. Impact on nonsense mediated decay transcript processing. Possible impact on downstream genes |
| c.1447_1451del | p.S483Pfs*5 | Intronic variant, possible impact on transcript processing |
| c.680- 681delAT |  | Intronic variant, possible impact on transcript processing |
| c.973-?_1081+?del | p.V325Tfs*7 | Intronic variant, possible retained intron. Could have impact on both upstream, downstream genes and nonsense medicated decay transcript processing. |
| c.972+3A>G |  | Intronic, splice region variant with low impact. Possible retained intron and impact on nonsense mediated decay transcript processing |
| c.326C>A | p.A109D | Missense variant with moderate impact, possible 5'UTR variant. Probably damaging to protein structure and deleterious to protein function. |
| c.336C>A | p.H112Q | Missense variant with moderate impact, possible 5'UTR variant. Probably damaging to protein structure and deleterious to protein function. |
| c.336C>G | p.H112Q | Missense variant with moderate impact, possible 5'UTR variant. Probably damaging to protein structure and deleterious to protein function. |
| c.336G>C | p.H112Q | Missense variant with moderate impact, possible 5'UTR variant. Probably damaging to protein structure and deleterious to protein function. |
| c.962G>A | p.G321E | Missense variant with moderate impact. Probably damaging to protein structure and deleterious to protein function |
| c.133C>T | p.A45T | Missense variant with moderate impact. Probably damaging to protein structure and deleterious to protein function. |
| c.1358A>G | p.Y453C | Missense variant with moderate impact. Probably damaging to protein structure and deleterious to protein function. |
| c.385A>C | p.T129P | Missense variant with moderate impact. Probably damaging to protein structure and deleterious to protein function. |
| c.932T>G | p.L311R | Missense variant with moderate impact. Probably damaging to protein structure and deleterious to protein function. Pathogenic. |
| c.1352T>G | p.L451W | Missense variant with potential impact on downstream gene regulation, possible 3'UTR variant involved in nonsense-mediated decay. Probably damaging to protein structure and likely deleterious to protein function. |
| c.1353G>T | p.L451F | Missense variant with potential impact on downstream gene regulation, possible 3'UTR variant involved in nonsense-mediated decay. Probably damaging to protein structure and likely deleterious to protein function. |
| c.1360G>C |  | Missense variant with potential impact on downstream gene regulation, possible 3'UTR variant involved in nonsense-mediated decay. Probably damaging to protein structure and likely deleterious to protein function. |
| c.1373T>A | p.V458D | Missense variant with potential impact on downstream gene regulation, possible 3'UTR variant involved in nonsense-mediated decay. Probably damaging to protein structure and likely deleterious to protein function. |
| c.1223G>A | p.C408Y | Missense variant with potential impact on downstream gene regulation, possible 3'UTR variant involved in nonsense-mediated decay. Probably damaging to protein structure and likely deleterious to protein function. |
| c.1348G>T | p.G450C | Missense variant with potential impact on downstream gene regulation, possible 3'UTR variant involved in nonsense-mediated decay. Probably damaging to protein structure and likely deleterious to protein function. |
| c.1367A>G | p.Y456C | Missense variant with potential impact on downstream gene regulation, possible 3'UTR variant involved in nonsense-mediated decay. Probably damaging to protein structure and likely deleterious to protein function.Pathogenic |
| c.1065C>A | p.F355L | Missense variant with potential impact on upstream and downstream gene regulation, possible 3'UTR variant involved in nonsense-mediated decay. Possible impact on processing of pseudogene. Benign protein structure and tolerated by protein function. |
| c.1052T>A | p.L351Q | Missense variant with potential impact on upstream and downstream gene regulation, possible 3'UTR variant involved in nonsense-mediated decay. Possible impact on processing of pseudogene. Probably damaging to protein structure and likely deleterious to protein function. |
| c.1057T>C | p.Y353H | Missense variant with potential impact on upstream and downstream gene regulation, possible 3'UTR variant involved in nonsense-mediated decay. Possible impact on processing of pseudogene. Probably damaging to protein structure and likely deleterious to protein function. |
| c.1069G>A | p.A357T | Missense variant with potential impact on upstream and downstream gene regulation, possible 3'UTR variant involved in nonsense-mediated decay. Possible impact on processing of pseudogene. Probably damaging to protein structure and likely deleterious to protein function. |
| c.1072G>A | p.G358R | Missense variant with potential impact on upstream and downstream gene regulation, possible 3'UTR variant involved in nonsense-mediated decay. Possible impact on processing of pseudogene. Probably damaging to protein structure and likely deleterious to protein function. |
| c.1078A>G | p.T360A | Missense variant with potential impact on upstream and downstream gene regulation, possible 3'UTR variant involved in nonsense-mediated decay. Possible impact on processing of pseudogene. Probably damaging to protein structure and likely deleterious to protein function. |
| c.140G>C | p.G47A | Missense variant, moderate impact and potential modifier of downstream gene regulation. Probably damaging to protein structure and likely to have deleterious effect on protein function |
| c.278T>C | p.I93T | Missense variant, moderate impact and potential modifier of downstream gene regulation. Probably damaging to protein structure and likely to have deleterious effect on protein function |
| c.506C>T | p.R169Q | Missense variant, moderate impact and potential modifier of downstream gene regulation. Probably damaging to protein structure and likely to have deleterious effect on protein function. |
| c.506G>A | R169Q | Missense variant, moderate impact and potential modifier of downstream gene regulation. Probably damaging to protein structure and likely to have deleterious effect on protein function. |
| c.533T>C | p.F178S | Missense variant, moderate impact and potential modifier of downstream gene regulation. Probably damaging to protein structure and likely to have deleterious effect on protein function. Possible retained intron. |
| c.139G>T | p.G47W | Missense variant, moderate impact and potential modifier of upstream and downstream gene regulation, possible impact on processing of pseudogene (FAM32BP). Probably damaging to protein structure and likely to have deleterious effect on protein function.Pathogenic |
| c.563T>C | p.L188P | Missense variant, moderate impact and potential modifier of upstream and downstream gene regulation. Probably damaging to protein structure and likely to have deleterious effect on protein function |
| c.578C>T | p.P193L | Missense variant, moderate impact and potential modifier of upstream and downstream gene regulation. Probably damaging to protein structure and likely to have deleterious effect on protein function. |
| c.139G>A | p.G47R | Missense variant, moderate impact and potential modifier of upstream and downstream gene regulation. Probably damaging to protein structure and likely to have deleterious effect on protein function. Conflicting evidence on clinical significance |
| c.650T>A | p.V217D | Missense variant, moderate impact and potential modifier of upstream gene regulation. Probably damaging to protein structure and likely to have deleterious effect on protein function |
| c.712G>A | p.D238N | Missense variant, moderate impact and potential modifier of upstream gene regulation. Probably damaging to protein structure and likely to have deleterious effect on protein function |
| c.872C>T | p.S291L | Missense variant, moderate impact and potential modifier of upstream gene regulation. Probably damaging to protein structure and likely to have deleterious effect on protein function. |
| c.620T>C |  | Missense variant, moderate impact and potential modifier of upstream gene regulation. Probably damaging to protein structure and likely to have deleterious effect on protein function. Uncertain clinical significance |
| c.791G>C | p.W264S | Missense variant, moderate impact and potential modifier of upstream gene regulation. Probably damaging/benign to protein structure and could have deleterious/tolerated impact on protein function. May cause retained intron. |
| c.1110C>A | p.N370K | Missense variant, moderate impact, possible 3'UTR variant involved in nonsense mediated decay. Probably damaging to protein structure and likely to have deleterious effect on protein function. |
| c.1445A>G |  | Missense variant, splice variant with potential impact on downstream gene regulation, possible 3'UTR variant involved in nonsense-mediated decay. Probably damaging to protein structure and likely deleterious to protein function. |
| c.1226C>A |  | Missense variant, splice variant with potential impact on downstream gene regulation, possible 3'UTR variant involved in nonsense-mediated decay. Probably damaging to protein structure and likely deleterious to protein function. |
| c.752C>T | p.P251L | Missense variant, splice region variant with moderate impact. Possibly damaging to protein structure and tolerated by protein function. |
| c.424G>A | p.G142S | Missense variant. Change tolerated by protein function, benign impact on protein structure |
| c.25C>T | p.R9W | Missense variant. Deleterious (but some evidnec of low confidence in finding) to protein function, benign impact on protein structure |
| c.2T>C | p.M1T | Missense variant. Deleterious (but some evidnec of low confidence in finding) to protein function, benign impact on protein structure |
| c.73G>T | p.G25C | Missense, splice region variant. Change tolerated by protein function and has benign impact on protein structure |
| c.882 -2A>G |  | Splice acceptor variant, high impact . Also potential impact on upstream gene regulation |
| c.973 -1G>A |  | Splice acceptor variant, high impact . Also potential regulatory region variant altering TF binding site. Could impact upstream gene regulation (RPL32P5) |
| c.973 -2A>G |  | Splice acceptor variant, high impact, may result in retained intron, could impact nonsense mediated decay . Also potential regulatory region variant altering TF binding site. Could impact upstream and downstream gene regulation (RPL32P5) |
| c.973-2A>G |  | Splice acceptor variant, high impact, may result in retained intron, could impact nonsense mediated decay . Also potential regulatory region variant altering TF binding site. Could impact upstream and downstream gene regulation (RPL32P5). |
| c.542+1G>A |  | Splice donor variant with high impact. Possible impact on nonsense mediated decay |
| c.753+2T>A |  | Splice donor variant with high impact. Possible retained intron |
| c.753G>A |  | Splice region variant with low impact. May influence downstream and upstream gene regulation. |
| c.781delinsCCATA | p.D261Pfs*2 | Stop gained, frameshift variant with high impact |
| c.1196G>A | p.W399* | Stop gained, high impact |
| c.794C>G | p.Q265X | Stop gained, high impact variant. Possible impact on upstream gene regulation |
| c.916C>T | p.R306* | Stop gained, high impact variant. Possible impact on upstream gene regulation. |
| c.660C>A | p.Y220X | Stop gained, high impact variant. Possible impact on upstream gene regulation. Benign. |
| c.47+2T>C |  | Synonymous, intron variant with low impact. Potential retained intron |

**Table IV C. *COL4A1***

| **Genetic mutation** | **Protein change** | **Variant information** |
| --- | --- | --- |
| c.*35C>A |  | 3' UTR variant, regulatory region variant |
| c.*31G>T |  | 3'UTR variant, regulatory region variant |
| c.*32G>A |  | 3'UTR variant, regulatory region variant |
| c.*32G>T |  | 3'UTR variant, regulatory region variant |
| c.*33T>A |  | 3'UTR variant, regulatory region variant |
| c.-2C>T |  | 5'UTR variant, with possible impact on upstream gene regulation |
| c.2545G>T | p.G808V | Evidence of stop gained, high impact |
| c.2424delT | p.P810fs | Frameshift mutation with high impact. Potentially leading to premature stop |
| c.2931dupT | p.G978WfsX15 | Frameshift mutation with high impact. Potentially leading to premature stop |
| c.3702delC | p. G1236* | Frameshift mutation with high impact. Potentially leading to premature stop |
| c.2085del | p.G696fs | Frameshift mutation with high impact. Potentially leading to premature stop. Pathogenic. |
| c.1121-18G>A |  | Intronic variant possibly leading to retained intron |
| c.2645_2646delinsAA | p.G882E | Intronic variant potentially leading to retained intron |
| c.3877-30C>A |  | Intronic variant with possible impact on upstream gene regulation. Intron retained |
| c.4582 -4586 dupCCCATG ins. |  | Intronic variant, retained intron. Likely deleterious and probably damaging. Possible impact on upstream gene regulation |
| c.4642T>G | p.C1548G | Missense & splice region variant with low to moderate effect. Likely to impact protein function and probably damaging |
| c.2969G>A | p.G990E | Missense variant and splice region variant. May result in retained intron. Possible modifier of downstream gene regulation. Likely deleterious and probably damaging. |
| c.2969G>T | p.G990V | Missense variant and splice region variant. May result in retained intron. Possible modifier of downstream gene regulation. Likely deleterious and probably damaging. |
| c.3200G>A | p. G1067E | Missense variant and splice region variant. May result in retained intron. Possible modifier of downstream gene regulation. Likely deleterious and probably damaging. |
| c.3200G>C | p.G1067A | Missense variant and splice region variant. May result in retained intron. Possible modifier of downstream gene regulation. Likely deleterious and probably damaging. |
| c.3770G>C | p.G1257E | Missense variant in possible regulatory region. Likely deleterious and probably damaging |
| c.3796G>C | p.G1266R | Missense variant in possible regulatory region. Likely deleterious and probably damaging |
| c.3832G>T | p.G1278S | Missense variant in possible regulatory region. Likely deleterious and probably damaging. Uncertain clinical significance |
| c.3245G>A | p.G1082E | Missense variant with moderate impact and possible modifier of downstream gene regulation. Likely deleterious and possibly damaging |
| c.3280G>C | p.G1094R | Missense variant with moderate impact and possible modifier of downstream gene regulation. Likely deleterious and possibly damaging |
| c.1249G>C | p.G417R | Missense variant with moderate impact. Benign impact on protein structure and deleterious to protein function |
| c.3997G>A | p.D1333N | Missense variant with moderate impact. Conflicting evidence of effect on protein function, potentially tolerated/potentially deleterious |
| c.3592G>A | p.G1198R | Missense variant with moderate impact. Likely deleterious and probably damaging |
| c.3620G>T | p.G1207V | Missense variant with moderate impact. Likely deleterious and probably damaging |
| c.3656G>A | p. G1219E | Missense variant with moderate impact. Likely deleterious and probably damaging |
| c.3671C>T | p.P1224L | Missense variant with moderate impact. Likely deleterious and probably damaging |
| c.3704A>G | p.K1235R | Missense variant with moderate impact. Likely deleterious and probably damaging |
| c.3706G>A | p.G1236R | Missense variant with moderate impact. Likely deleterious and probably damaging |
| c.3707G>A | p. G1237E | Missense variant with moderate impact. Likely deleterious and probably damaging |
| c.3712C>T | p.R1238C | Missense variant with moderate impact. Likely deleterious and probably damaging |
| c.3505G>A | p. G1169S | Missense variant with moderate impact. Possible splice region variant, with potential impact on downstream gene regulation. Likely deleterious and probably damaging |
| c.2512A>G | p.M838V | Missense variant with moderate impact. Possibly damaging to protein structure and deleterious to protein function |
| c.3389G>A | p.G1130D | Missense variant with moderate impact. Potentially modifies upstream and downstream gene regulation. Likely deleterious and probably damaging |
| c.4088 G > A | p.G1363D | Missense variant with moderate impact. Probably damaging and deleterious to protein function |
| c.1502G>A |  | Missense variant with moderate impact. Probably damaging to protein structure and deleterious to protein function |
| c.1528G>A | p.G510R | Missense variant with moderate impact. Probably damaging to protein structure and deleterious to protein function |
| c.1583G>A | p.G528E | Missense variant with moderate impact. Probably damaging to protein structure and deleterious to protein function |
| c.1619A>G | p.K540R | Missense variant with moderate impact. Probably damaging to protein structure and deleterious to protein function |
| c.2008G>A | p.G670R | Missense variant with moderate impact. Probably damaging to protein structure and deleterious to protein function |
| c.2045G>T | p. G682V | Missense variant with moderate impact. Probably damaging to protein structure and deleterious to protein function |
| c.2063G>A | p.G688D | Missense variant with moderate impact. Probably damaging to protein structure and deleterious to protein function |
| c.2078G>A | p.G693E | Missense variant with moderate impact. Probably damaging to protein structure and deleterious to protein function |
| c.2086G>A | p.G696S | Missense variant with moderate impact. Probably damaging to protein structure and deleterious to protein function |
| c.2086G>T | p.G696C | Missense variant with moderate impact. Probably damaging to protein structure and deleterious to protein function |
| c.2132G>A | p.G711E | Missense variant with moderate impact. Probably damaging to protein structure and deleterious to protein function |
| c.2159G>A | p.G720D | Missense variant with moderate impact. Probably damaging to protein structure and deleterious to protein function |
| c.2168G>A | p. G723E | Missense variant with moderate impact. Probably damaging to protein structure and deleterious to protein function |
| c.2504G>A | p.G835E | Missense variant with moderate impact. Probably damaging to protein structure and deleterious to protein function |
| c.625G>A | p. G209S | Missense variant with moderate impact. Probably damaging to protein structure and deleterious to protein function |
| c.634G>A | p.G212S | Missense variant with moderate impact. Probably damaging to protein structure and deleterious to protein function |
| c.1493G>A | p.G498D | Missense variant with moderate impact. Probably damaging to protein structure and deleterious to protein function. Pathogenic. |
| c.1493G>T | p.G498V | Missense variant with moderate impact. Probably damaging to protein structure and deleterious to protein function. Pathogenic. |
| c.3383T>A | p.I1128N | Missense variant with moderate impact. Substitution seems to be tolerated by protein function but probably damaging to protein structure |
| c.3715G>A | p.G1239R | Missense variant with possible impact on upstream gene regulation. Probably damaging variant |
| c.3941G>T | p.G1314V | Missense variant with possible impact on upstream gene regulation. Probably damaging variant |
| c.3976G>A | p.G1326R | Missense variant with possible impact on upstream gene regulation. Probably damaging variant |
| c.3995G>A | p.G1332D | Missense variant with possible impact on upstream gene regulation. Probably damaging variant |
| c.4031G>C | p.G1344A | Missense variant with possible impact on upstream gene regulation. Probably damaging variant |
| c.4105G>C | p.G1369R | Missense variant with possible impact on upstream gene regulation. Probably damaging variant |
| c.4213G>A | p.G1405S | Missense variant with possible impact on upstream gene regulation. Probably damaging variant |
| c.1801G>A | p. G601S | Missense variant, moderate impact and potential modifier of downstream gene regulation. Only possibly damaging to protein structure and likely to have deleterious effect on protein function |
| c.1807C>T | p.P603S | Missense variant, moderate impact and potential modifier of downstream gene regulation. Only possibly damaging to protein structure and likely to have deleterious effect on protein function |
| c.1555G>A | p.G519R | Missense variant, moderate impact and potential modifier of downstream gene regulation. Probably damaging to protein structure and likely to have deleterious effect on protein function |
| c.1835G>A | p.G612D | Missense variant, moderate impact and potential modifier of downstream gene regulation. Probably damaging to protein structure and likely to have deleterious effect on protein function |
| c.1853G > A | p.G618E | Missense variant, moderate impact and potential modifier of downstream gene regulation. Probably damaging to protein structure and likely to have deleterious effect on protein function |
| c.2494G>A | p.G832R | Missense variant, moderate impact and potential modifier of downstream gene regulation. Probably damaging to protein structure and likely to have deleterious effect on protein function |
| c.2563G>C | p.G855R | Missense variant, moderate impact and potential modifier of downstream gene regulation. Probably damaging to protein structure and likely to have deleterious effect on protein function |
| c.2581G>A | p.G861S | Missense variant, moderate impact and potential modifier of downstream gene regulation. Probably damaging to protein structure and likely to have deleterious effect on protein function |
| c.2599G>A | p.G867R | Missense variant, moderate impact and potential modifier of downstream gene regulation. Probably damaging to protein structure and likely to have deleterious effect on protein function |
| c.2608G>A | p.G870R | Missense variant, moderate impact and potential modifier of downstream gene regulation. Probably damaging to protein structure and likely to have deleterious effect on protein function |
| c.2636G>A | p.G879E | Missense variant, moderate impact and potential modifier of downstream gene regulation. Probably damaging to protein structure and likely to have deleterious effect on protein function |
| c.2645G>A | p.G882D | Missense variant, moderate impact and potential modifier of downstream gene regulation. Probably damaging to protein structure and likely to have deleterious effect on protein function |
| c.2662G>A | p.G888R | Missense variant, moderate impact and potential modifier of downstream gene regulation. Probably damaging to protein structure and likely to have deleterious effect on protein function |
| c.2689G>A | p.G897S | Missense variant, moderate impact and potential modifier of downstream gene regulation. Probably damaging to protein structure and likely to have deleterious effect on protein function |
| c.2699G>A | p.G900E | Missense variant, moderate impact and potential modifier of downstream gene regulation. Probably damaging to protein structure and likely to have deleterious effect on protein function |
| c.2744G>A | p.G915E | Missense variant, moderate impact and potential modifier of downstream gene regulation. Probably damaging to protein structure and likely to have deleterious effect on protein function |
| c.2782G>C | p.D928H | Missense variant, moderate impact and potential modifier of downstream gene regulation. Probably damaging to protein structure and likely to have deleterious effect on protein function |
| c.2842G>A | p.G948S | Missense variant, moderate impact and potential modifier of downstream gene regulation. Probably damaging to protein structure and likely to have deleterious effect on protein function |
| c.2987G>A | p.G996D | Missense variant, moderate impact and potential modifier of downstream gene regulation. Probably damaging to protein structure and likely to have deleterious effect on protein function |
| c.3022G>A | p.G1008R | Missense variant, moderate impact and potential modifier of downstream gene regulation. Probably damaging to protein structure and likely to have deleterious effect on protein function |
| c.3040G>C | p.G1014R | Missense variant, moderate impact and potential modifier of downstream gene regulation. Probably damaging to protein structure and likely to have deleterious effect on protein function |
| c.3104G>T | p.G1035V | Missense variant, moderate impact and potential modifier of downstream gene regulation. Probably damaging to protein structure and likely to have deleterious effect on protein function |
| c.3122G>A | p.G1041E | Missense variant, moderate impact and potential modifier of downstream gene regulation. Probably damaging to protein structure and likely to have deleterious effect on protein function |
| c.3130G>C | p.G1044E | Missense variant, moderate impact and potential modifier of downstream gene regulation. Probably damaging to protein structure and likely to have deleterious effect on protein function |
| c.3190G>A | p.G1064S | Missense variant, moderate impact and potential modifier of downstream gene regulation. Probably damaging to protein structure and likely to have deleterious effect on protein function |
| c.191G>T | p.G64V | Missense variant, moderate impact and potential modifier of upstream gene regulation. Probably damaging to protein structure and likely to have deleterious effect on protein function. Possible 3'UTR variant. |
| c.4739G>C | p.G1580A | Missense variant, moderate impact, deleterious and likely to impact protein function, probably damaging |
| c.4881C>G | p.N1627K | Missense variant, moderate impact, deleterious and likely to impact protein function, probably damaging |
| c.4843G>A | p.E1615K | Missense variant, Moderate impact, possibly retained intron, probably damaging |
| c.4232G>C | p.G1411A | Missense variant, moderate impact. Probably damaging and likely to have deleterious effect on protein function |
| c.4380T>G | p.C1460W | Missense variant, moderate impact. Probably damaging and likely to have deleterious effect on protein function |
| c.4652G>A | p. C1551Y | Missense variant, moderate impact. Probably damaging and likely to have deleterious effect on protein function |
| c.4717G>A | p.G1573R | Missense variant, moderate impact. Probably damaging and likely to have deleterious effect on protein function |
| c.4738 G > A | p.G1580S | Missense variant, moderate impact. Probably damaging and likely to have deleterious effect on protein function |
| c.4738G>A | p. G1580S | Missense variant, moderate impact. Probably damaging and likely to have deleterious effect on protein function |
| c.1955G>A | p. G652E | Missense variant, moderate impact. Probably damaging to protein structure and likely to have deleterious effect on protein function |
| c.1963G>A | p.G655R | Missense variant, moderate impact. Probably damaging to protein structure and likely to have deleterious effect on protein function |
| c.1964G>A | p.G655E | Missense variant, moderate impact. Probably damaging to protein structure and likely to have deleterious effect on protein function |
| c.1973C>A | p. G658V | Missense variant, moderate impact. Probably damaging to protein structure and likely to have deleterious effect on protein function |
| c.1973G>A | p.G658D | Missense variant, moderate impact. Probably damaging to protein structure and likely to have deleterious effect on protein function |
| c.2441 G > T | p.G814V | Missense variant, non-coding exon variant with moderate impact. Possibly damaging to protein structure and deleterious to protein function |
| c.2413G>A | p.G805R | Missense variant, non-coding exon variant with moderate impact. Possibly damaging to protein structure and deleterious to protein function |
| c.2413G>C | p. G805R | Missense variant, non-coding exon variant with moderate impact. Possibly damaging to protein structure and deleterious to protein function |
| c.2317G>A | p.G773R | Missense variant, non-coding exon variant with moderate impact. Probably damaging to protein structure and deleterious to protein function |
| c.2317G>C | p.G773R | Missense variant, non-coding exon variant with moderate impact. Probably damaging to protein structure and deleterious to protein function |
| c.2228G>T | p.G743V | Missense variant, non-coding exon variant with moderate impact. Regulatory region variant, leading to open chromatin structure. Probably damaging to protein structure and deleterious to protein function |
| c.2245G>A | p.G749S | Missense variant, non-coding exon variant with moderate impact. Regulatory region variant, leading to open chromatin structure. Probably damaging to protein structure and deleterious to protein function |
| c.2263G>A | p.G755R | Missense variant, non-coding exon variant with moderate impact. Regulatory region variant, leading to open chromatin structure. Probably damaging to protein structure and deleterious to protein function |
| c.4267G>C | p.G1423R | Missense variant, possibly resulting in retained intron. Possibly damaging and likely deleterious to protein function |
| c.4133G>A | p.G1378D | Missense variant, potentially impacting upstream gene regulation. Likely deleterious and probably damaging |
| c.4150+1(IVS46) G>T |  | Missense variant, splice donor variant with potential impact on upstream gene regulation. High impact. Probably damaging and likely deleterious. |
| c.4150+1G>A |  | Missense variant, splice donor variant with potential impact on upstream gene regulation. High impact. Probably damaging and likely deleterious. |
| c.4150G>A | p.G1384S | Missense variant, splice donor variant with potential impact on upstream gene regulation. High impact. Probably damaging and likely deleterious. |
| c.2345G>C | p.G782A | Missense variant, splice region variant with low-moderate impact. Likely deleterious and probably damaging |
| c.2096G>A | pG699D | Missense variant, splice region variant with low-moderate impact. Likely deleterious to protein function and probably damaging to protein structure |
| c.236G>T | p.G79V | Missense variant, splice region variant with low-moderate impact. Likely deleterious to protein function and probably damaging to protein structure |
| c.443G>A | p.G148E | Missense variant, splice region variant with low-moderate impact. Likely deleterious to protein function and probably damaging to protein structure |
| c.196C>A | p.Q66K | Missense variant. Change tolerated by protein function but possibly damaging to protein structure |
| c.2641A>G | p.M881V | Missense variant. Change tolerated by protein function but possibly damaging to protein structure |
| c.3046A>G | p.M1016V | Missense variant. Change tolerated by protein function but possibly damaging to protein structure |
| c.31C>A | p.L11M | Missense variant. Change tolerated by protein function but possibly damaging to protein structure |
| c.1612C>G | p.R538G | Missense variant. Change tolerated by protein function with benign impact on protein structure |
| c.1769G>A | p.G562E | Missense variant. Change tolerated by protein function with benign impact on protein structure |
| c.3946C>G | p.Q1316E | Missense variant. Change tolerated by protein function, likely benign some evidence of possibly damaging protein structure |
| c.1537-2A>G |  | Potential frameshift variant and splice acceptor variant with high impact |
| c.1537–2delA |  | Potential frameshift variant and splice acceptor variant with high impact |
| c.1121-2dupA | p.G374_N429 delinsD | Splice acceptor variant, intronic variant leading to retained intron. High impact variant. |
| c.1382-1G>C |  | Splice acceptor variant, intronic variant leading to retained intron. High impact variant. |
| c.2194-1G.A |  | Splice acceptor variant, intronic variant leading to retained intron. High impact variant. Also potential regulatory region variant leading to open chromatin structure |
| c.553-2A>G |  | Splice acceptor variant, intronic variant leading to retained intron. High impact variant. Also potential regulatory region variant leading to open chromatin structure and altered downstream gene regulation. Potential 3' UTR variant |
| c.1990+1G>A |  | Splice donor variant with high impact. Possible retained intron |
| c.3406 + 1G>T |  | Splice donor variant with high impact. Potential impact on both upstream and downstream gene regulation |
| c.2716 + 1G>A |  | Splice donor variant with high impact. Potential impact on downstream gene regulation |
| c.2716+ G>T |  | Splice donor variant with high impact. Potential impact on downstream gene regulation |
| c.2716+2T>C |  | Splice donor variant with high impact. Potential impact on downstream gene regulation |
| c.2458+1G>A |  | Splice donor variant, high impact. Possibly retained intron and downstream gene regulation modification |
| c.1A>T |  | Start lost, but seems to be tolerated by protein function but possibly damaging to protein structure. Possible impact on upstream gene regulation |
| c.739C>T | p.Q247* | Stop gained, high impact. Possible modifier of downstream gene regulation |
| c.607G>T | p. G203R | Stop gained, high impact. Potential 3'UTR regulatory variant |
| c.4875C>A | p.Y1625* | Stop gained, likely deleterious, high impact |
| c.4887C>A | p.Y1629X | Stop gained, likely deleterious, high impact |
| c.1870G>T | p.G624* | Stop gained, likely deleterious, high impact. Possible modifier of downstream gene regulation |

**Table IV D. *COL4A2***

| **Genetic mutation** | **Protein change** | **Variant information** |
| --- | --- | --- |
| c.1396G>A | p.G466S | Missense variant with moderate impact. Probably damaging to protein structure and deleterious to protein function. Possible intron variant causing alteration to lncRNA influencing gene AS2 |
| c.1776+1G>A |  | Splice donor variant with high impact. Possible retained intron and impact to lncRNA influencing gene AS2. Pathogenic but also reported to have uncertain clinical significance |
| c.1810G>C | p.G604R | Missense variant, moderate impact and potential modifier of upstream and downstream gene regulation. Probably damaging to protein structure and likely to have deleterious effect on protein function. Potenital influence on promoter refulation and lncRNA influencing AS2 |
| c.1856G>A | p.G619D | Missense variant, moderate impact and potential modifier of upstream and downstream gene regulation. Probably damaging to protein structure and likely to have deleterious effect on protein function. Potenital influence on promoter refulation and lncRNA influencing AS2. Likely pathogenic |
| c.2105G>A | p.G702D | Missense variant with moderate impact. Probably damaging to protein structure and deleterious to protein function |
| c.2399G>A | p.G800E | Missense variant with moderate impact. Probably damaging to protein structure and deleterious to protein function. With possible impact on upstream gene regulation and promoter regions |
| c.2821G>A | p.G941R | Missense variant, non-coding exon variant with moderate impact. Possibly damaging to protein structure and deleterious to protein function. Possible retained intron. |
| c.3110G>A | p.G1037E | Missense variant, moderate impact and potential modifier of downstream gene regulation. Probably damaging to protein structure and likely to have deleterious effect on protein function. Pathogenic |
| c.3368A>G | p.E1123G | Missense variant, non-coding exon variant with moderate impact. Possibly damaging to protein structure and deleterious to protein function. Likely benign clinical significance but possible risk factor |
| c.3448C>A | p.Q1150K | Missense variant, non-coding exon variant with moderate impact. Possibly damaging to protein structure and tolerated to protein function. Likely benign clinical significance but possible risk factor |
| c.3455G>A | p.G1152D | Missense variant, splice region variant. Probably damaging to protein structure and deleterious to protein function. Pathogenic |
| c.3490G>A | p.R1164G | Missense variant with moderate impact. Probably damaging to protein structure and deleterious to protein function. Pathogenic |
| c.4129G > A | p.G1377R | Missense variant, moderate impact and potential modifier of upstream gene regulation, possible impact on lncRNA influencing AS2. Probably damaging to protein structure and likely to have deleterious effect on protein function. Pathogenic |
| c.4147G>A | p.G1383R | Missense variant, moderate impact and potential modifier of upstream gene regulation, possible impact on lncRNA influencing AS2. Probably damaging to protein structure and likely to have deleterious effect on protein function. Likely pathogenic |
| c.4987G>A | p.G1663S | Missense variant, moderate impact and potential modifier of both upstream and downstream gene regulation, possible impact on lncRNA influencing AS2. Probably damaging to protein structure and likely to have deleterious effect on protein function. Conflicting clinical significane, reported both likely benign and likely pathogenic |
| c.5068G>A | p.A1690T | Missense variant, non-coding exon variant with moderate impact. Possibly damaging to protein structure and tolerated to protein function. Likely benign clinical significance but possible risk factor |

**Table IV E. *TREX1***

| **Genetic mutation** | **Protein change** | **Variant information** |
| --- | --- | --- |
| c.703dup | p.V235GfsX6 | Frameshift mutation with high impact. Potentially leading to premature stop. Possible impact on downstream gene regulation of ATRIP and SHISA5 (non-mediated decay) |
| c.822delT | p.P275Qfsx2 | Frameshift mutation with high impact. Potentially leading to premature stop. Possible impact on downstream gene regulation of ATRIP and SHISA5 (non-mediated decay) |
| c.830-833dupAGGA | p.D278fs | Intronic variant. Potential 3'UTR variant with downstream gene variation. Possible influence on ATRIP and nonsense mediated decay of SHISA5 |
| c.829A>T | p.K277* | Stop gained, high impact. Possible modifier of downstream gene regulation. Likely pathogenic. |
| c.828_831dupGAAG | p.D278EfsTer48 | Frameshift mutation with high impact. Potentially leading to premature stop. Possible impact on downstream gene regulation of ATRIP and SHISA5 (non-mediated decay) |
| c.703dupG | p.V235Gfs | Frameshift mutation with high impact. Potentially leading to premature stop. Possible impact on downstream gene regulation of ATRIP and SHISA5 (non-mediated decay) |
| c.685A>G | p.Arg229Gly | Missense variant, moderate impact and potential modifier of downstream gene regulation. Benign impact on protein structure and tolerated by protein function |
| c.690G>T | p.Lys230Asn | Missense variant, moderate impact and potential modifier of downstream gene regulation. Benign impact on protein structure and could have deleterious effect on protein function, but tolerated also reported |
| c.581delC | p.Ala194fs | Frameshift variant with high impact, possible downstream gene regulation of ATRIP and SHISA5. Pathogenic |
| c.742_745dupGTCA | p.T249fs | Intronic variant. Potential 3'UTR variant with downstream gene variation. Possible influence on ATRIP and nonsense mediated decay of SHISA5 |
| c.734dupC | ? | Frameshift mutation with high impact. Potentially leading to premature stop. Possible impact on downstream gene regulation of ATRIP and SHISA5 (non-mediated decay) |
| c.911_912delCA | p.T304Nfs*12 | Intronic variant. Potential 3'UTR variant with downstream gene variation. Possible influence on ATRIP and nonsense mediated decay of SHISA5 |
| c.703_704insG | p.V235GfsX6 | Frameshift mutation with high impact. Potentially leading to premature stop. Possible impact on downstream gene regulation of ATRIP and SHISA5 (non-mediated decay) |

**Appendix I. Search Strategy**

1. CADASIL/

2. (CADASIL or "Cerebral autosomal dominant arterio$ with subcortical infarct$ and leukoencephalopathy" or (Dementia and hereditary and multi?infarct) or "Familial vascular leukoencephalopathy" or CASIL or "Cerebral arterio$ with subcortical infarct$ and leukoencephalopathy" or "Chronic familial vascular encephalopathy" or "Familial disorder with subcortical ischemic stroke$" or "Agnogenic medial arteriopathy" or "Familial Binswanger$ disease" or (cerebral and autosomal dominant and arterio$ and infarct$ and leukoencephalophy)).af.

3. (CARASIL or "Maeda$ syndrome" or "Cerebral autosomal recessive arterio$ with subcortical infarct$ and leukoencephalopathy" or ("Subcortical Vascular Encephalopathy" and Progressive) or "Cerebrovascular Disease With Thin Skin Alopecia And Disc Disease" or "Nemoto disease" or (cerebral and autosomal recessive and arterio$ and infarct$ and leukoencephalophy) or "Familial young adult onset arterio$ leukoencephalopathy with alopecia and lumbago").af.

4. ((COL4A1$ and (leukoencephalopathy or small vessel disease or autosomal dominant or infantile hemiparesis or retinal arter$ tortuosity or RATOR or PADMAL or "pontine autosomal dominant microangiopathy and leukoencephalopathy" or Walker Warburg or porencephaly 1 or "small vessel disease of the brain with or without ocular abnormalities" or BSVD)) or HANAC or (hereditary angio$ and nephropath$ and aneurysm$ and cramp$) or ((autosomal dominant or familial or hereditary) and (h?ematuria and Retinal Arter$ Tortuosity)) or ("Autosomal dominant familial porencephaly" or "Hereditary multi infarct dementia" or HEMID or hMID) or (multi-infarct dementia and Swedish) or "Nonsyndromic autosomal dominant congenital cataract").af.

5. Muscle Cramp/ and Raynaud Disease/

6. (COL4A2 and (Porencephaly or stroke or Microbleed$ or h?emorrhage or leukoencephalopathy or small vessel disease or autosomal recessive or infantile hemiparesis or retinal arter$ tortuosity)).af.

7. (RVCL or "Retinal vasculopathy with cerebral leukodystrophy" or ($retinal vascul$ and (hereditary or familial)) or ((Cerebroretinal Vasculopathy and Hereditary) or "hereditary vascular retinopathy") or "Grand-Kaine-Fulling syndrome" or HERNS or Hereditary Systemic Angiopathy or (hereditary and endotheliopathy and retin$ and nephro$ and stroke$) or (hereditary and retin$ and (raynaud$ or migraine)) or ADRVCL or (Autosomal Dominant and Retin$ and (leukodystrophy or leukoenchalopathy))).af.

8. ("Early-onset stroke and vasculopathy associated with mutations in ADA2" or (Stroke and vasc$ and ADA2) or ((deficien$ and (ADA 2 or ADA2 or adenosine deaminase-2)) or DADA2 or DADA 2 or (Vasculitis and ADA2 deficien$)) or Sneddon Syndrome or (Polyarteritis nodosa and Childhood onset)).af.

9. (CARASAL or (Cathepsin A related arteriopathy with stroke? and leukoencephalopathy)).af.

10. 1 or 2 or 3 or 4 or 5 or 6 or 7 or 8 or 9
11. (NOTCH?3 or Notch 3 or "Neurogenic locus notch homolog protein 3").af.

12. (TREX?1 or TREX 1 or "Three prime repair exonuclease 1").af.

13. (COL4A1 or COL4A2 or COL4 A1 or COL4 A2 or "COL4 A 1" or "COL4 A 2" or "COL 4 A1" or "COL 4 A2").af.

14. (Collagen and ("type IV" or "type 4") and (alpha?1 or alpha?2 or alpha 1 or alpha 2)).af.

15. Collagen Type IV/

16. (alpha?1 or alpha?2 or alpha 1 or alpha 2).af.

17. 15 and 16

18. (HTRA?1 or HTRA 1 or "HtrA serine peptidase 1" or "HtrA serine protease 1").af.

19. (CECR?1 or CECR 1 or "Cat eye syndrome critical region protein 1" or "adenosine deaminase 2" or ADA2 or ADA 2).af.

20. (FOXC?1 or FOX C1 or FOXC 1 or "FOX C 1" or "forkhead box C?1" or "Forkhead box C 1").af.

21. (PITX?2 or PITX 2 or "paired-like homeodomain 2" or "pituitary homeobox 2" or "Paired-like homeodomain transcription factor 2").af.

22. (Cathepsin?A or Cathepsin A or CathA or Cath A or CTSA).af.

23. 11 or 12 or 13 or 14 or 17 or 18 or 19 or 20 or 21 or 22

24. exp Cerebral Small Vessel Diseases/

25. exp Cerebrovascular Disorders/

26. exp stroke/

27. exp dementia, vascular/

28. Brain Diseases/

29. exp basal ganglia cerebrovascular disease/

30. exp brain ischemia/

31. exp intracranial arterial diseases/

32. exp Cerebral Hemorrhage/

33. exp intracranial hemorrhages/

34. leukomalacia, periventricular/

35. stroke, lacunar/

36. Leukoaraiosis/

37. Leukoencephalopathies/

38. White Matter/

39. Infarction/

40. ("Cerebral Small Vessel Disease?" or cerebrovascular).af.

41. (White matter hyperintensit$ or WMH$ or White matter MR hyperintensit$ or White matter magnetic resonance hyperintensit$ or Subcortical hyperintensit$ or White matter lesion? or WML$ or Hyper intensit$ or Leukodystroph$ or Leukoaraiosis or Leukomalacia or White Matter Change? or WMC? or White Matter Disease or WMD or White matter damage or Grey matter hyperintensit$ or Brainstem hyperintensit$ or Subcortical hyperintensit$ or White matter hypoattenuation? or White matter hypodensit$ or Leukoencephalopath$).af.

42. (Subcortical infarct? or Cerebral infarct$ or Brain infarct$ or Silent brain infarct$ or Striatocapsular infarct$ or Lacunar infarct$ or Lacune? or Lacunar stroke? or Lacunar syndrome or Stroke? or Vascular lesion?).af.

43. (Microbleed? or Cerebral Microbleed or CMB? or Hypointense lesion? or Subcortical H?emorrhage or Intracerebral h?emorrhage or Cortical siderosis or Superficial siderosis).af.

44. (Perivascular space? or Virchow Robin space? or Type 3 lacune? or Etat crible).af.

45. (Brain atrophy or Cerebral atrophy or Global atrophy or Corpus callosum atrophy or Central atrophy or Mesencephalic atrophy or Hippocampal atrophy or Cortical thinning).af.

46. 24 or 25 or 26 or 27 or 28 or 29 or 30 or 31 or 32 or 33 or 34 or 35 or 36 or 37 or 38 or 39 or 40 or 41 or 42 or 43 or 44 or 45

47. 23 and 46
48. 10 or 47

49. limit 48 to humans

50. remove duplicates from 49
